# Increased Insulin Resistance and Hyperglycemia in Long COVID disease: A Systematic Review and Meta-Analysis

**DOI:** 10.1101/2025.11.01.25339290

**Authors:** Abbas F. Almulla, Yingqian Zhang, Chavit Tunvirachaisakul, Andre F Carvalho, Michael Maes

**Author notes:** **Corresponding authors:** Prof. Dr. Michael Maes, and Yingqian Zhang., Sichuan Provincial Center for Mental Health, Sichuan Provincial People’s Hospital, School of Medicine, University of Electronic Science and Technology of China, Chengdu 610072, China.

## Abstract

**Background:** accumulating evidence suggests that Long COVID (LC) is mediated by chronic immune activation, oxidative stress, and metabolic dysregulation. These processes may impair glucose homeostasis and promote insulin resistance (IR). However, no prior meta-analysis has systematically and quantitatively evaluated IR indices and related biomarkers in LC compared with normal controls.

**Objectives:** To systematically review and meta-analyze composite and solitary indices of IR, β-cell function, and adipokine levels in individuals with LC compared with normal controls.

**Methods:** PubMed, SCOPUS, and Google Scholar databases were searched for relevant studies from inception to August 2025. Sixty-three eligible studies were included, comprising 12,409 participants—5,891 LC patients and 6,518 normal controls.

**Results:** LC disease is characterized by elevated global IR (standardized mean difference, SMD = 0.395; 95% confidence intervals, CI: 0.226;0.563), Fasting insulin + C-peptide + FBG composite score (SMD = 0.605; 95% CI: 0.306;0.904) and acute + chronic glycemia composite scores (SMD = 0.424; 95% CI: 0.258;0.590). Furthermore, significant increases in HOMA-IR (SMD = 0.621; 95% CI: 0.379;0.863), Insulin (SMD = 0.488; 95% CI: 0.202;0.774), HbA1c (SMD = 0.308; 95% CI: 0.053;0.563), and fasting and random blood glucose (SMD = 0.831; 95% CI: 0.271;1.391, SMD = 0.396; 95% CI: 0.188;0.605) alongside reduced HOMA-%B and HOMA-%S were observed in LC patients versus normal controls. No publication bias observed in the results.

**Conclusion:** The current study suggests that LC disease is characterized by persistent insulin resistance, hyperglycemia, and β-cell dysfunction, suggesting sustained metabolic disturbances beyond the acute phase

## Introduction

A majority of individuals infected with coronavirus disease (COVID-19) experience recovery; however, a subset may develop long COVID (LC) condition, which can result in medium- to long-term effects influencing one or more organ systems (World Health Organization 2025). The prevalence of this condition is reported to be as high as 36% and can reach up to 85% among hospitalized patients (Pavli, Theodoridou et al. 2021, Hou, Gu et al. 2025). Clinical manifestations are diverse and affect multiple systems, with over 200 reported symptoms, including cardiovascular complications, metabolic disturbances, pancreatic involvement, and frequent insulin resistance (IR) following the acute stage (Dennis, Wamil et al. 2021, Khunti, Davies et al. 2021, Al-Hakeim, Al-Rubaye et al. 2023).

The pathophysiological mechanisms underpinning LC are not fully elucidated and may encompass direct viral injury, immune dysregulation, oxidative stress, autoimmunity, atherogenicity, and prolonged organ damage (Almulla, Thipakorn et al. 2024a, Almulla, Thipakorn et al. 2024c, Almulla, Maes et al. 2024d, Almulla, Thipakorn et al. 2025). LC is marked by heightened immune-inflammatory responses alongside oxidative and nitrosative stress (Al-Hakeim, Al-Rubaye et al. 2023, Almulla, Thipakorn et al. 2024a). A recent meta-analysis reveals that elevated immune-response system, M1 macrophage activity, T helper 1 and Th17 responses, along with increased C-reactive protein concentrations, are significant characteristics (Almulla, Thipakorn et al. 2024a). Elevated atherogenic indices are noted, characterized by decreased high-density lipoprotein (HDL) and apolipoprotein (Apo) A levels, alongside increased low-density lipoprotein (LDL), total cholesterol, ApoB, and triglycerides (Almulla, Thipakorn et al. 2025). Importantly, IR is significantly linked to abnormal lipid metabolism, immune activation, and liver dysfunction which are all observed in LC (Avramoglu, Basciano et al. 2006, Cetin, Demir et al. 2020, Berbudi, Khairani et al. 2025).

Acute COVID-19 markedly affects glucose metabolism via various mechanisms, resulting in hyperglycemia and IR (Grubišić, Švitek et al. 2023). Severe acute respiratory syndrome coronavirus 2 (SARS-CoV-2) has a direct impact on pancreatic beta cells, essential for insulin synthesis (Wu, Lidsky et al. 2021). The virus attaches to ACE2 receptors in the pancreas and other metabolic organs, which may lead to beta-cell dysfunction and reduced insulin secretion (Kazakou, Lambadiari et al. 2022). Harding et al. show that COVID-19 survivors exhibit increased susceptibility to developing new-onset diabetes mellitus (DM) (Harding, Oviedo et al. 2023). Glycemic abnormalities can continue after recovery, with changes in glucose metabolism and IR observed for up to two months following infection (Montefusco, Ben Nasr et al. 2021, Al-Hakeim, Al-Rubaye et al. 2023). Additionally, numerous studies have indicated that LC is associated with increased fasting glucose, insulin levels, and Homeostasis Model Assessment for Insulin Resistance (HOMA-IR), alongside diminished insulin sensitivity (Al-Hakeim, Al-Rubaye et al. 2023, Al-Hakeim, Khairi Abed et al. 2023, Bota, Bratosin et al. 2024). IR is increasingly acknowledged as a factor in the clinical manifestations of LC including neuropsychiatric symptoms, such as fatigue, anxiety, and depressive symptoms (Al-Hakeim, Al-Rubaye et al. 2023, Al-Hakeim, Khairi Abed et al. 2023, Maes, Almulla et al. 2023).

The new-onset DM is likely attributable to direct injury to beta cells, systemic inflammation, and the use of glucocorticoids during treatment, which exacerbate hyperglycemia and IR (Grubišić, Švitek et al. 2023). Systemic inflammation in severe COVID-19 might worsen IR and hyperglycemia, with increased levels of interleukin (IL)-6, IL-2, IL-10, and IFN-γ associated with impaired glucose metabolism (Montefusco, Ben Nasr et al. 2021, Zheng, Wang et al. 2021). Oxidative and nitrosative stress and reduced antioxidant capacity, which are evident in LC, might impact glucose regulation (Soto, Guarner-Lans et al. 2022).

Although IR is frequently observed in LC, no previous meta-analysis has quantitatively evaluated IR indicators in LC patients compared with normal controls. Therefore, this study examines IR indices in individuals with LC relative to controls. Solitary biomarkers comprise HbA1c, fasting blood glucose (FBG), random blood sugar (RBS), HOMA-IR, HOMA-%B, HOMA-%S, insulin, and C-peptide, along with adipokines such as adiponectin, leptin, and ghrelin. Composite measures include global IR, fasting insulin + C-peptide + fasting glucose, and acute + chronic glycemia. We hypothesize that LC is associated with increased insulin resistance and hyperglycemia

## Materials and Methods

This meta-analysis was designed and conducted in alignment with the Preferred Reporting Items for Systematic Reviews and Meta-Analyses (PRISMA 2020) statement (Page, McKenzie et al. 2021). Methodological decisions were additionally guided by the Cochrane Handbook for Systematic Reviews of Interventions (Chandler, Cumpston et al. 2019) and the recommendations from the Meta-analysis of Observational Studies in Epidemiology (MOOSE).

The study population comprised individuals with LC compared to control groups. We evaluated a broad spectrum of IR and glucose metabolism markers, including HOMA-IR, HOMA-β%, HOMA-S%, fasting insulin, HbA1c, and C-peptide. To provide a more comprehensive assessment, circulating levels of adipokines—notably adiponectin, leptin, and ghrelin—were also examined. In addition to single biomarker analysis, we computed several composite indices that integrate multiple parameters of IR and β-cell function. These included the Global IR index, fasting insulin + C-peptide + fasting glucose, and the acute + chronic glycemia index.

This systematic review and meta-analysis was prospectively registered in the PROSPERO database under registration number CRD42025637597.

### Search strategy

A systematic literature search was performed to identify studies reporting biomarkers of IR and glycemia in LC patients. The search included primary electronic databases, namely PubMed/MEDLINE, Google Scholar, and SCOPUS, spanning from inception to September 2025. Search queries were formulated using a combination of predefined keywords and Medical Subject Headings (MeSH) terms, as detailed in Table 1 of the Electronic Supplementary File (ESF). We also conducted a manual review of reference lists from eligible articles and relevant meta-analyses, thereby minimizing the risk of missing pertinent publications.

**Table 1.**
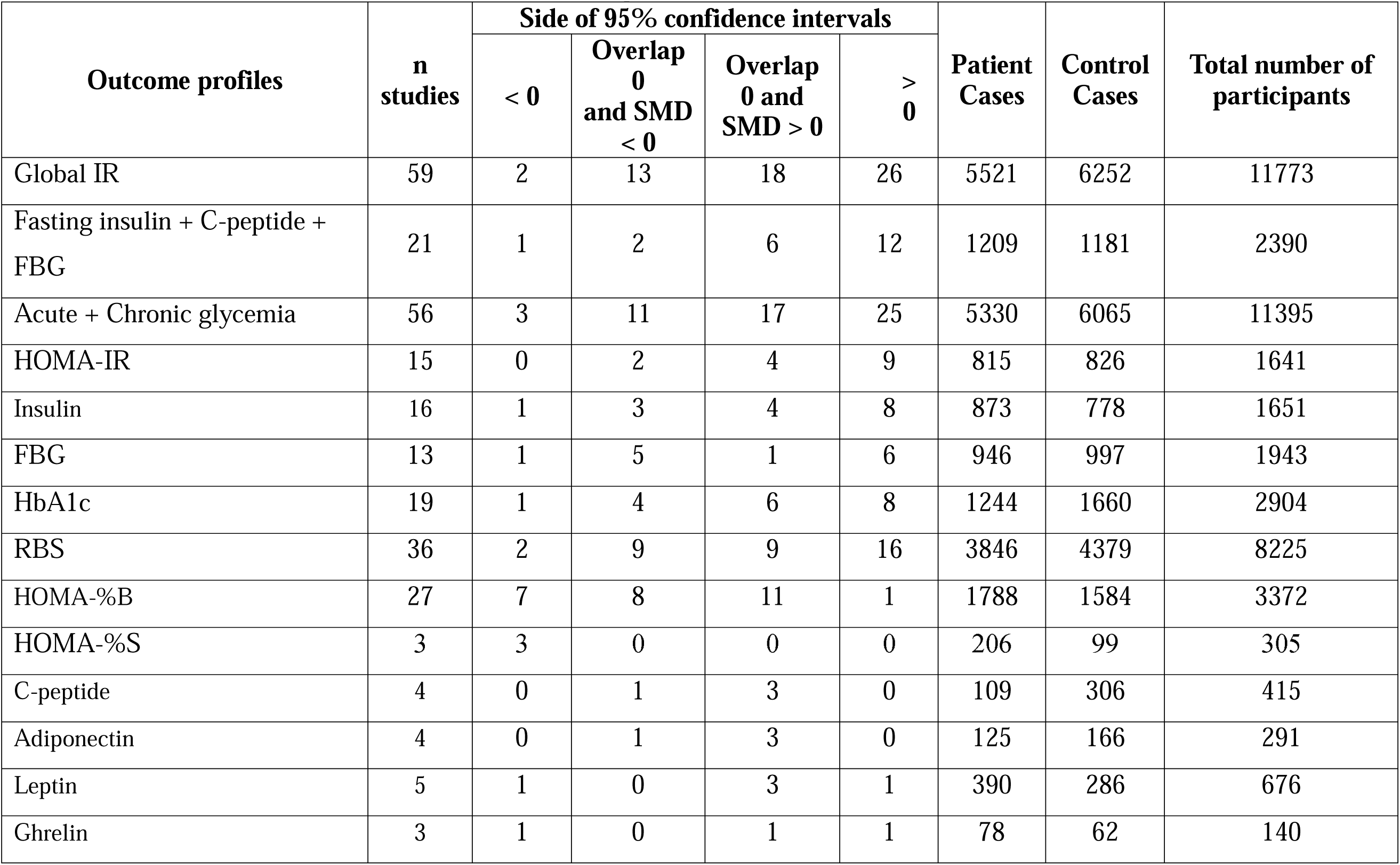
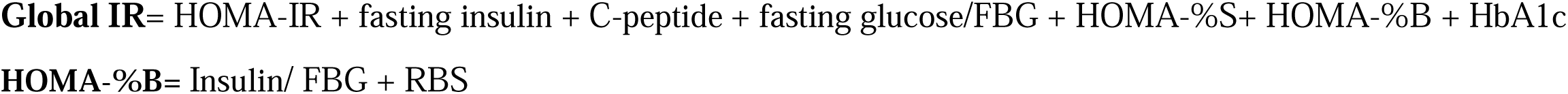
The outcomes and number of patients with Long COVID (LC) and normal controls along with the side of standardized mean difference (SMD) and the 95% confidence intervals with respect to zero SMD.

### Eligibility criteria

A thorough search was conducted to identify eligible studies, prioritizing peer-reviewed publications in English. However, grey literature and reports in other languages, such as Thai, French, Spanish, Turkish, Hungarian, German, Ukrainian, Italian, Russian, and Arabic, were also included in the consideration. As in our previous meta-analysis, studies were included if they utilized an observational design (case–control or cohort) with a control group, examined IR biomarkers in patients with LC diagnosed per WHO criteria (2021) (World Health Organization 2021), and offered baseline biomarker assessments alongside longitudinal follow-up.

The exclusion criteria included studies utilizing unconventional biological materials, such as hair, saliva, platelet-rich plasma, or cerebrospinal fluid; studies limited to genetic or translational analyses; studies that did not include control groups; and studies that lacked data on means with standard deviation (SD) or standard error (SE), unless these values were obtainable from the authors or could be calculated from published data using validated tools like the online calculator (hkbu.edu.hk, https://automeris.io/WebPlotDigitizer/).

### Primary and secondary outcomes

This meta-analysis assessed various biomarkers indicative of IR, emphasizing composite indices such as the Global IR, fasting insulin + C-peptide + fasting glucose, and the acute + chronic glycemia index, in addition to the main IR biomarkers including HOMA-IR, insulin, and FBG as detailed in **Table 1**. Alongside the integrated measures, secondary analyses examined individual biomarkers including HbA1c, RBS, HOMA-%B, HOMA-%S, and C-peptide. Levels of adipokines, specifically adiponectin, leptin, and ghrelin, were also evaluated.

### Screening and data extraction

Two investigators (AA and YZ) independently performed screening and data extraction, initially evaluating study titles and abstracts based on our predefined eligibility criteria. Full texts of potentially relevant articles were acquired for thorough evaluation, and studies that fulfilled the exclusion criteria were eliminated. Key information was systematically organized into a structured Excel sheet, including study authors, publication year, biomarker values for IR (mean and standard deviation), participant numbers in both patient and control groups, and total sample size. Recorded variables encompassed study design, biological sample type (serum, plasma, blood), duration since acute infection, intensive care unit admission during the initial illness, participant age, sex distribution, and geographic setting. Discrepancies in data entry were addressed in consultation with the senior author (MM). The studies’ methodological quality was assessed using the Immunological Confounder Scale (ICS), first introduced by Andrés-Rodríguez et al. (2020) and later adapted by MM for IR research in LC (Andrés-Rodríguez, Borràs et al. 2020). The ICS consists of two complementary instruments: the Quality Scale, which evaluates study design characteristics such as sample size, confounder control, and sampling duration (scoring range 0–10, with higher scores indicating superior quality), and the Redpoints Scale, which assesses the degree of potential bias in LC immune activation research (scoring range 0–26, with lower scores reflecting greater methodological rigor). These instruments have been employed in prior meta-analyses related to immune activation and kynurenine pathway disruptions in LC, as well as in studies on lipid peroxidation and tryptophan metabolism in affective disorders (Almulla, Thipakorn et al. 2022a, Almulla, Thipakorn et al. 2023, Almulla, Thipakorn et al. 2024, Almulla, Thipakorn et al. 2024a).

### Data analysis

In the present meta-analysis, we employed Comprehensive Meta-Analysis (CMA) software, version 4, in line with PRISMA recommendations (see ESF, Table 2) to conduct all analyses. Biomarkers were included in the analysis only when at least two independent studies provided data. To generate a composite estimate of global IR, we integrated values from HOMA-IR + fasting insulin + C-peptide + fasting glucose + HbA1c + RBS + HOMA-2S, considering their interdependence. The Fasting insulin + C-peptide + fasting glucose index was calculated by combining, fasting insulin, C-peptide, fasting glucose under the same dependency assumption. Likewise, the Acute + Chronic glycemia was derived by integrating HbA1c, FBG and RBS values. Pooled effect sizes were computed using a random-effects model with constrained maximum likelihood estimation, with results expressed as standardized mean differences (SMDs) and 95% confidence intervals (CIs). A two-tailed p-value < 0.05 was considered statistically significant. Interpretation of effect sizes followed Cohen’s guidelines, where SMD values of 0.20, 0.50, and 0.80 correspond to small, medium, and large effects, respectively. Heterogeneity was assessed through τ², Q statistics, and I², while meta-regression analyses were applied to examine moderators of variability. Robustness of results was tested using leave-one-out sensitivity analyses. To evaluate publication bias, we applied the fail-safe N test, continuity-corrected Kendall’s tau, and Egger’s regression intercept (one-tailed for the latter two). Where Egger’s test indicated asymmetry, the trim-and-fill method was employed to estimate missing studies and adjust pooled outcomes. Funnel plots were generated to visually examine the relationship between study precision and SMD, including both observed and imputed studies. Meta-regression analyses were performed to investigate potential sources of heterogeneity. Moderators including age, sex, latitude, and post-COVID duration were individually incorporated into the regression models. Results were presented as z-values alongside their respective one-tailed p-values, derived from a random-effects model framework.

**Table 2.**
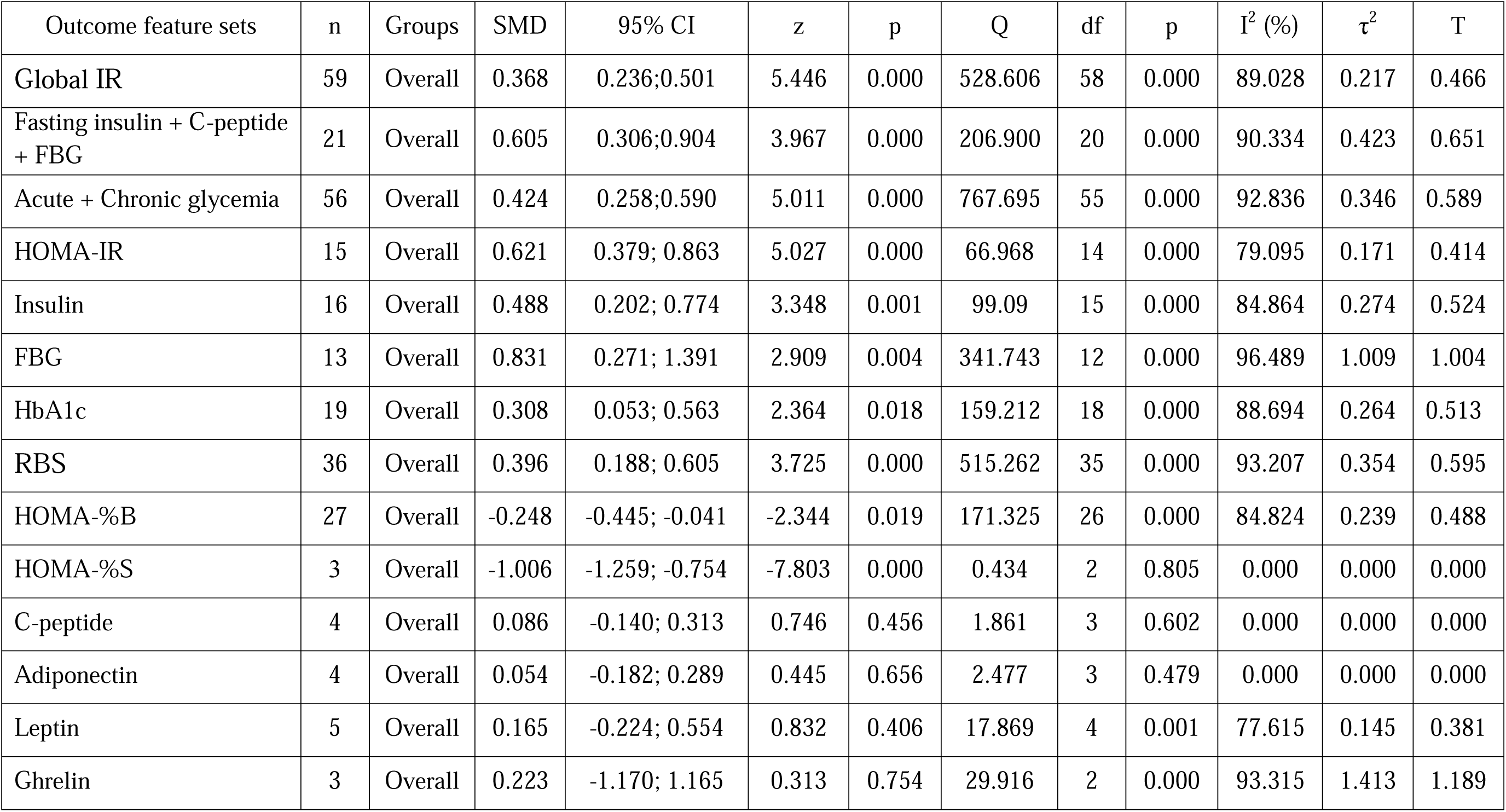
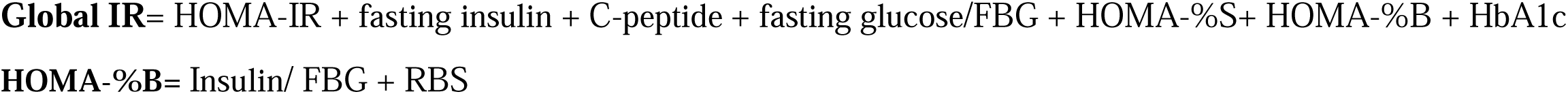
Results of meta-analysis performed on several outcomes (atherogenic indices and lipid profile) variables with combined different media and separately.

## Results

### Search Outcomes

Databases including PubMed, Google Scholar, SciFinder, and SCOPUS were systematically searched using prespecified keywords and MeSH terms (see ESF Table 1). As shown in the PRISMA flowchart (**Figure 1**), the initial search yielded 1253 articles. After removing duplicates and irrelevant studies, 82 remained for detailed screening. Of these, 63 met the inclusion criteria for the systematic review, and 63 studies were ultimately included in the meta-analysis according to the predefined eligibility standards (Holmes, Wist et al. 2021, Jud, Gressenberger et al. 2021, Mittal, Ghosh et al. 2021, Montefusco, Ben Nasr et al. 2021, Vimercati, De Maria et al. 2021, Орлова, Ломайчиков et al. 2021, Alfadda, Rafiullah et al. 2022, Clemente, Sinatti et al. 2022, Duran, Kurtipek et al. 2022, Keerthi, Sushmita et al. 2022, Labarca, Henríquez-Beltrán et al. 2022, Meisinger, Goßlau et al. 2022, Sumbalová, Kucharská et al. 2022, Tong, Yan et al. 2022, Verma, Ramayya et al. 2022, Abdulaziz Alsufyani 2023, Adatsi, Bockarie et al. 2023, Al-Hakeim, Al-Rubaye et al. 2023, Al-Hakeim, Khairi Abed et al. 2023, Al Masoodi, Radhi et al. 2023, Alshehri, AlQahtani et al. 2023, Erol, Tezcan et al. 2023, López-Hernández, Oropeza-Valdez et al. 2023, Mora, Kogut et al. 2023, Paris, Palomba et al. 2023, Rajamanickam, Venkataraman et al. 2023, Silva, Pereira et al. 2023, Szczerbiński, Okruszko et al. 2023, Tudoran, Bende et al. 2023, Vyas, Joshi et al. 2023, Xuereb, Borg et al. 2023, Yamamoto, Otsuka et al. 2023, Ach, Ben Haj Slama et al. 2024, Agafonova, Elovikova et al. 2024, Al-Zadjali, Al-Lawati et al. 2024, Atieh, Durieux et al. 2024, Bielecka-Dabrowa, Kapusta et al. 2024, Bota, Bratosin et al. 2024, Di Ciaula, Liberale et al. 2024, Domingo, Battistini et al. 2024, Duarte, Sambra et al. 2024, Emiroglu, Dicle et al. 2024, Fernandez-de-las-Peñas, Notarte et al. 2024, Ghosh, Niesen et al. 2024, Gupta, Nicholas et al. 2024, Inceu, Nechifor et al. 2024, Kartika, Subekti et al. 2024, Korkmaz, Çınar et al. 2024, Kuryata, Mytrokhina et al. 2024, Kuryłowicz, Babicki et al. 2024, Manuilov and Mykhailovska 2024, Matviichuk, Yerokhovych et al. 2024, Matviichuk, Yerokhovych et al. 2024, Oliván-Blázquez, Bona-Otal et al. 2024, Parás-Bravo, Fernández-de-Las-Peñas et al. 2024, Struttmann, Shah et al. 2024, Torki, Hoseininasab et al. 2024, Vojdani, Almulla et al. 2024, Zerón-Rugerio, Zaragozá et al. 2024, Al Masoodi, Radhi et al. 2025, Barichello, Kluwe-Schiavon et al. 2025, Jamwal, Chhabra et al. 2025, Visconti, Rocha et al. 2025).

**Figure 1.**
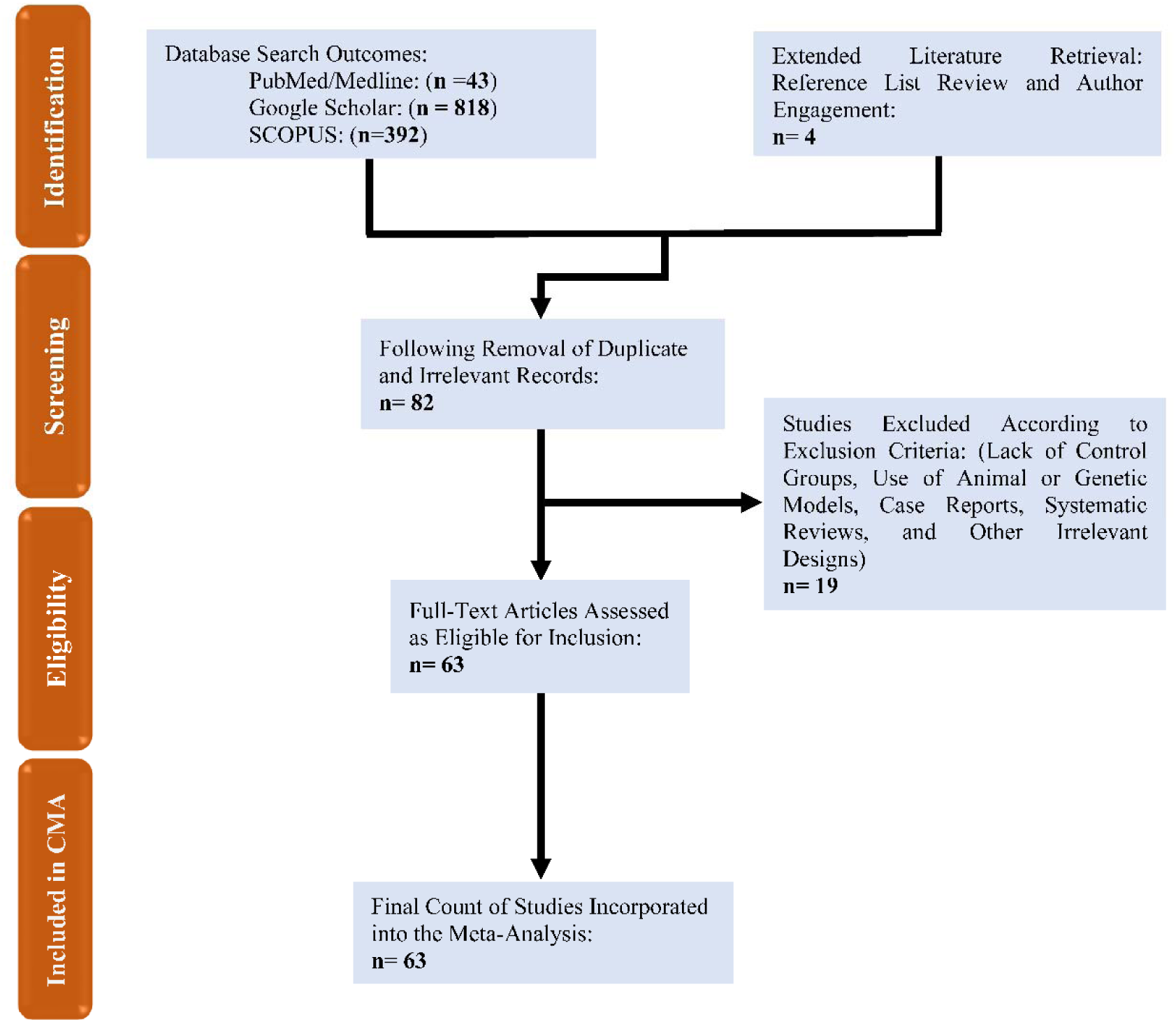
The PRISMA flow chart.

The meta-analysis comprised 12409 participants, including 5891 LC patients and 6518 control subjects, with ages ranging from 11 to 70 years. Most studies utilized routine laboratory tests alongside ELISA techniques to assess IR biomarkers. The included studies represented a wide geographic distribution, led by United State (20) and Italy (5). Iraq, Spain, and Turkey each contributed 4 studies, while Saudi Arabia contributed 3 studies. Brazil, India, Poland, Romania, Russia, and Ukraine each provided 2 studies. Additional single-study contributions were from Australia, Chile, China, Germany, Indonesia, Iran, Japan, Malta, Oman, Slovakia, and UK. Study quality assessment showed median (min–max) values of 4 (min = 0, max = 9.5) for quality and 14.5 (min = 3.5, max = 26) for redpoints (ESF Table 3).

**Table 3.** Results on publication bias.

| Outcome feature sets | Fail safe n | Z Kendall's $\tau$ | p | Egger's t test (df) | p | Missing studies (side) | After Adjusting |
| --- | --- | --- | --- | --- | --- | --- | --- |
| Global IR | 14.221 | 0.954 | 0.169 | 1.551(57) | 0.063 | 10 (Right) | - |
| Fasting insulin + C-peptide + fasting glucose/FBG | 11.381 | 0.150 | 0.439 | 1.600(19) | 0.063 | 0 | - |
| Acute + Chronic glycemia | 14.719 | 1.208 | 0.113 | 1.767(54) | 0.041 | 13 (Right) | 0.683 (0.496;0.870) |
| HOMA-IR | 11.457 | 0.890 | 0.186 | 1.339 (13) | 0.101 | 0 | - |
| Insulin | 7.8/52 | 0.045 | 0.482 | 1.431(14) | 0.087 | 0 | - |
| FBG | 12.593 | 1.403 | 0.080 | 1.617(11) | 0.067 | 2 (Right) | - |
| HbA1c | 7.467 | 0.209 | 0.416 | 0.656(17) | 0.260 | 2 (Right) | - |
| RBS | 11.098 | 1.266 | 0.102 | 1.391(34) | 0.086 | 9 (Right) | - |
| HOMA-%B | -5.373 | 1.292 | 0.098 | 1.027(25) | 0.156 | 0 | - |
| HOMA-%S | -7.795 | 0.000 | 0.500 | 0.305(1) | 0.405 | 0 | - |
| C-peptide | 1.211 | 1.019 | 0.154 | 5.359 (2) | 0.016 | 2 (Left) | 0.013 (-0.192; 0.220) |
| Adiponectin | 0.319 | 0.339 | 0.367 | 0.568 (2) | 0.313 | 1 (Left) | - |
| Leptin | 1.882 | 0.244 | 0.403 | 0.134 (3) | 0.450 | 1 (Left) | 0.083 (-0.277; 0.445) |
| Ghrelin | 1.359 | 1.044 | 0.148 | 16.18(1) | 0.0196 | 0 | 0 |

**Global IR**= HOMA-IR + fasting insulin + C-peptide + fasting glucose/FBG + HOMA-%S+ HOMA-%B + HbA1c
**HOMA-%B**= Insulin/ FBG + RBS

## Primary Outcome Variables

### Global IR

Thirty-six studies evaluated the global IR composite (Table 1, **Figure 2**). Only one study had CIs entirely below zero, 15 entirely above, and 20 overlapping (7 negative, 13 positive SMDs). LC patients showed a significant increase in global IR (**Table 2**, Figure 2). No publication bias was found (**Table 3**).

**Figure 2.**
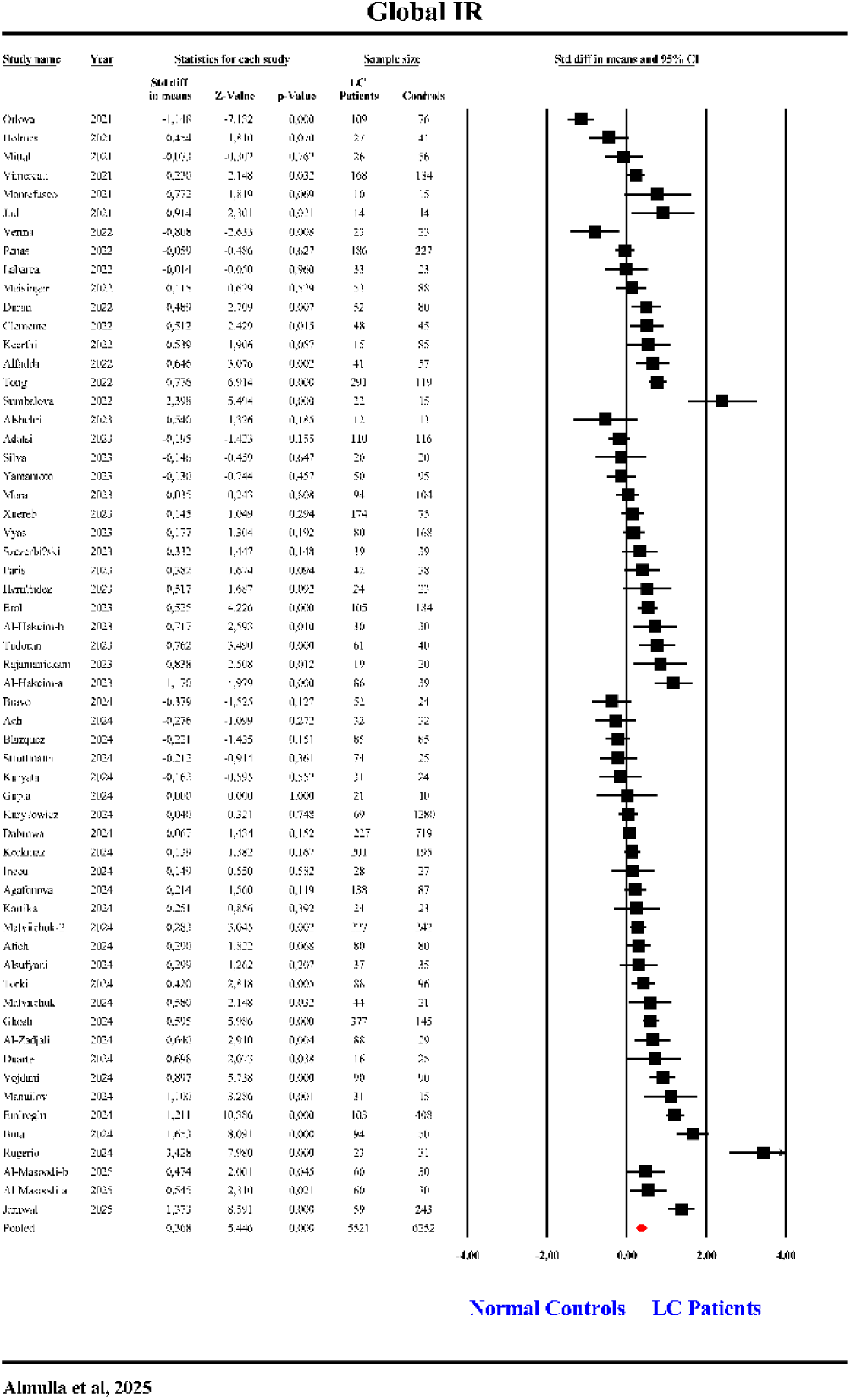
The forest plot of global insulin resistance (IR) in patients with Long COVID (LC) and normal controls.

### Fasting Insulin + C-Peptide + Fasting Glucose

The meta-analysis included 21 studies evaluating the composite of fasting insulin, C-peptide, and fasting glucose (Table 1, **Figure 3**). One study presented CIs entirely below zero, while 12 studies had CIs entirely above zero. Eight studies showed overlapping CIs, with 2 reporting negative and 6 reporting positive SMDs. Overall, LC patients exhibited a significant increase in this IR composite compared to controls (Table 2, Figure 3). No publication bias was detected (Table 3).

**Figure 3.**
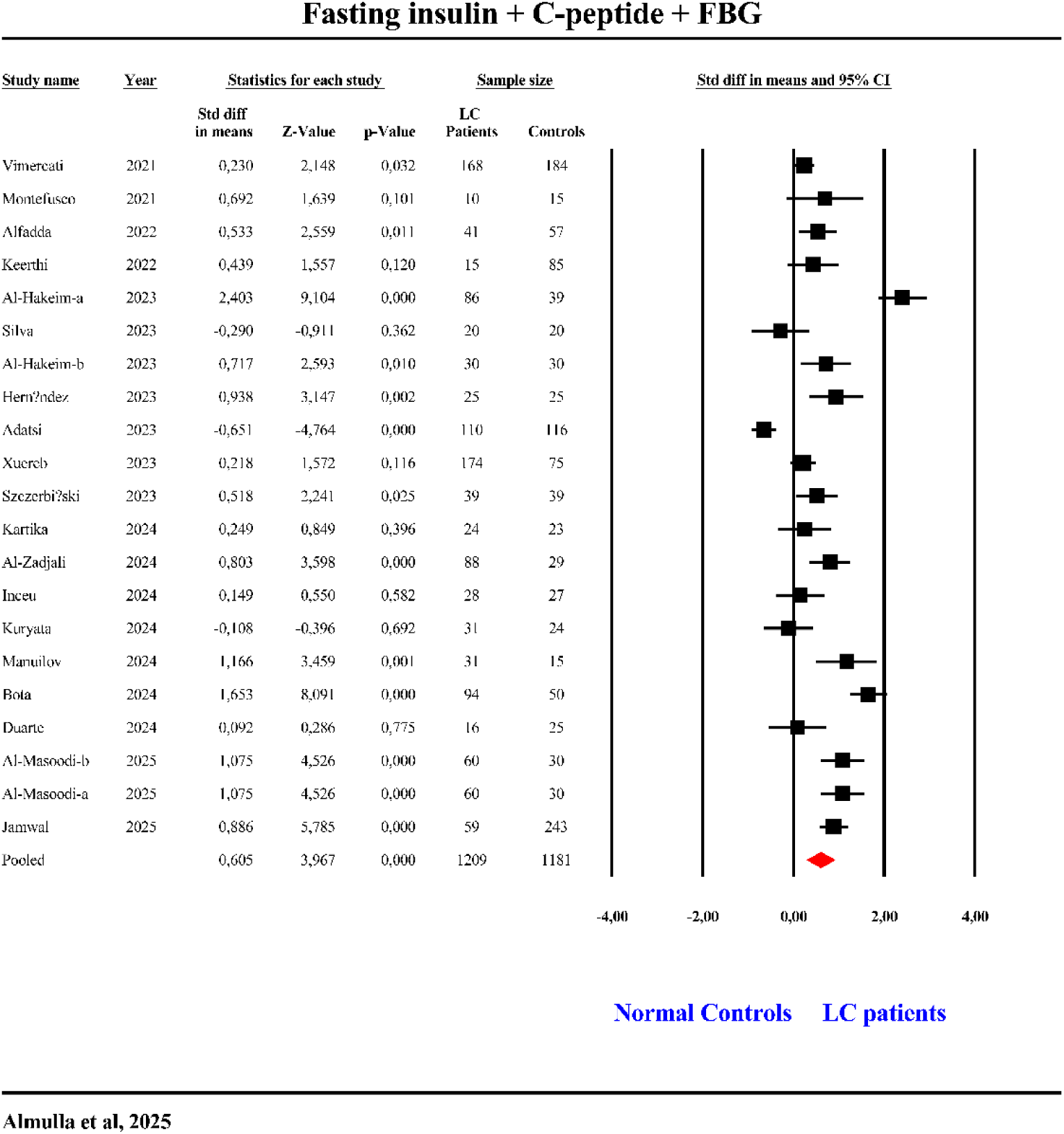
The forest plot of Fasting insulin + C-peptide + FBG in patients with Long COVID (LC) and normal controls.

### Acute + Chronic Glycemia

Fifty-six studies examined the acute + chronic glycemia composite (Table 1, **Figure 4**). Three studies had CIs entirely below zero, and 25 had CIs entirely above zero. Twenty-eight showed overlapping CIs (11 negative, 17 positive SMDs). The composite was significantly elevated in LC patients (Table 2, Figure 4). Funnel plot analysis revealed publication bias, with 13 missing studies on the right side; adjustment increased the pooled SMD to 0.683 (Table 3).

**Figure 4.**
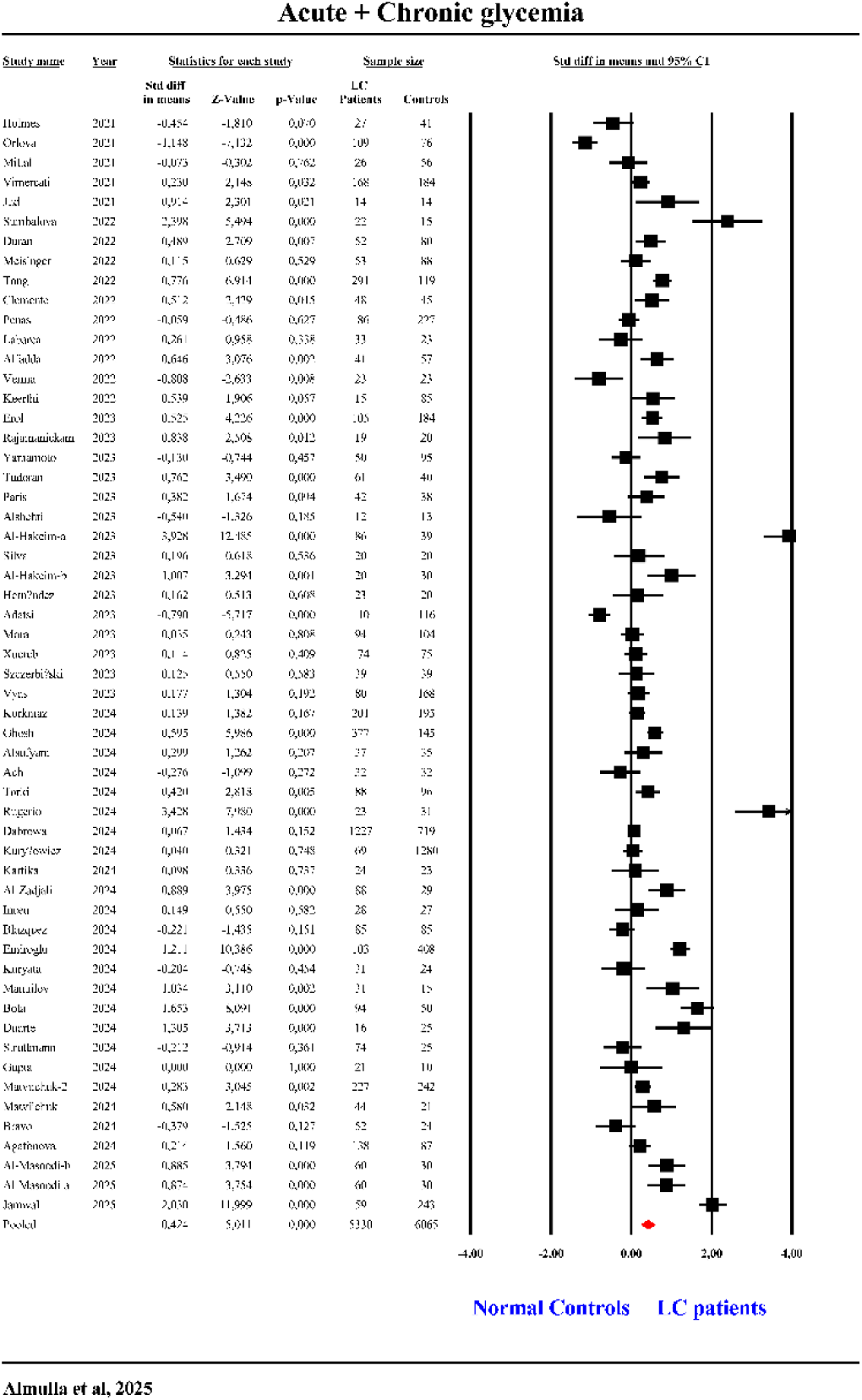
The forest plot of Acute + Chronic glycemia in patients with Long COVID (LC) and normal controls.

### HOMA-IR

Fifteen studies assessed HOMA-IR (Table 1, **Figure 5**). None reported CIs entirely below zero, while 9 had CIs entirely above. Six had overlapping CIs (2 negative, 4 positive SMDs). LC patients exhibited significantly higher HOMA-IR values, with a large effect size (SMD = 0.621) (Table 2, Figure 5). No publication bias was found (Table 3).

**Figure 5.**
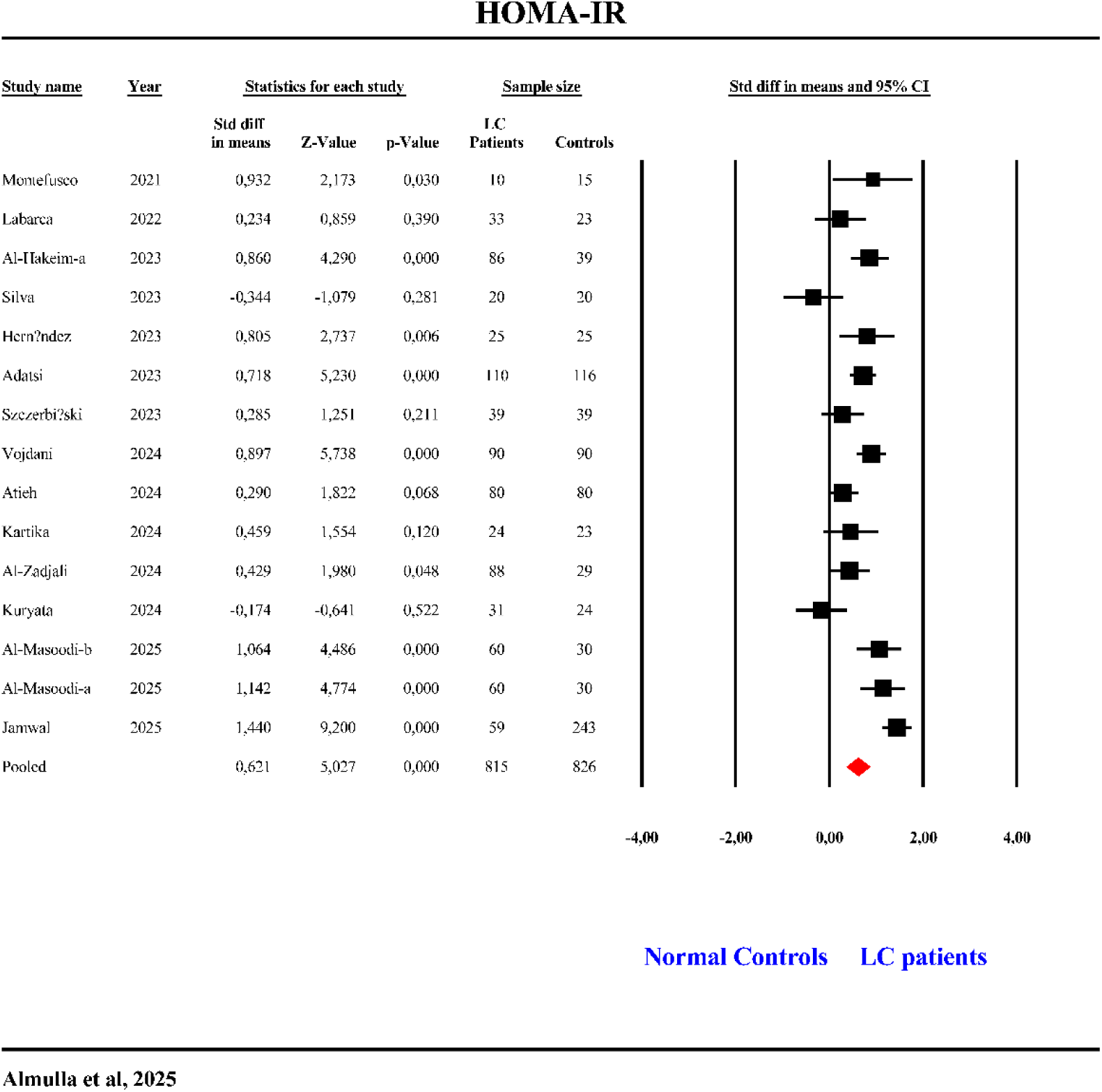
The forest plot of HOMA-IR in patients with Long COVID (LC) and normal controls.

### Insulin

Sixteen studies examined insulin levels (Table 1, **Figure 6**). One study reported CIs below zero, 8 above, and 7 overlapping (3 negative, 4 positive SMDs). LC patients showed significantly increased insulin levels (Table 2, **Figure 6**). Egger’s and Kendall tests confirmed no publication bias (Table 3).

**Figure 6.**
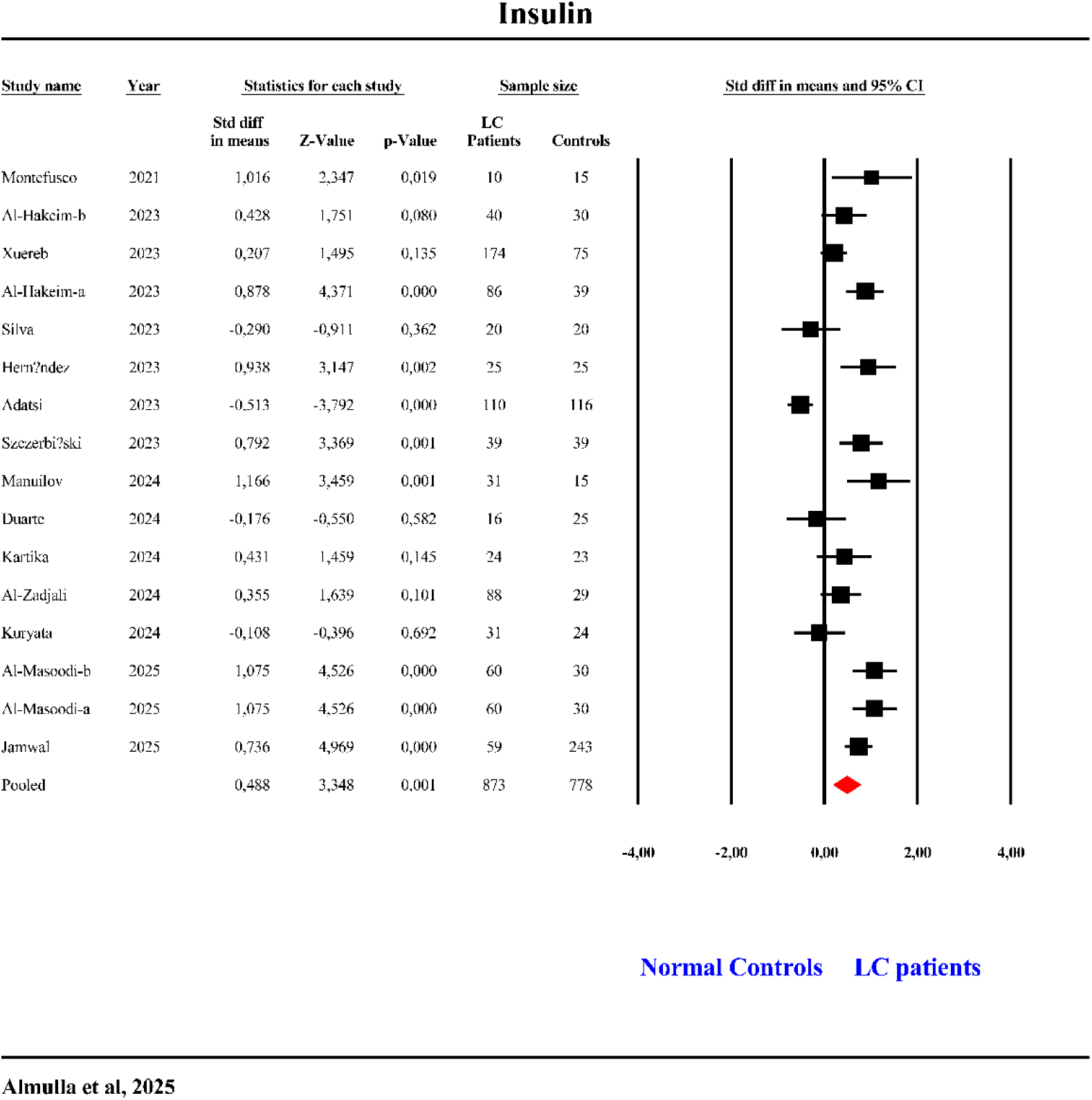
The forest plot of insulin in patients with Long COVID (LC) and normal controls.

### FBG

Thirteen studies assessed FBG (Table 1, **Figure 7**). One study had CIs below zero, 6 above, and 6 overlapping (1 negative, 5 positive SMDs). FBG levels were significantly elevated with a large effect size in LC patients (Table 2). No publication bias was found (Table 3).

**Figure 7.**
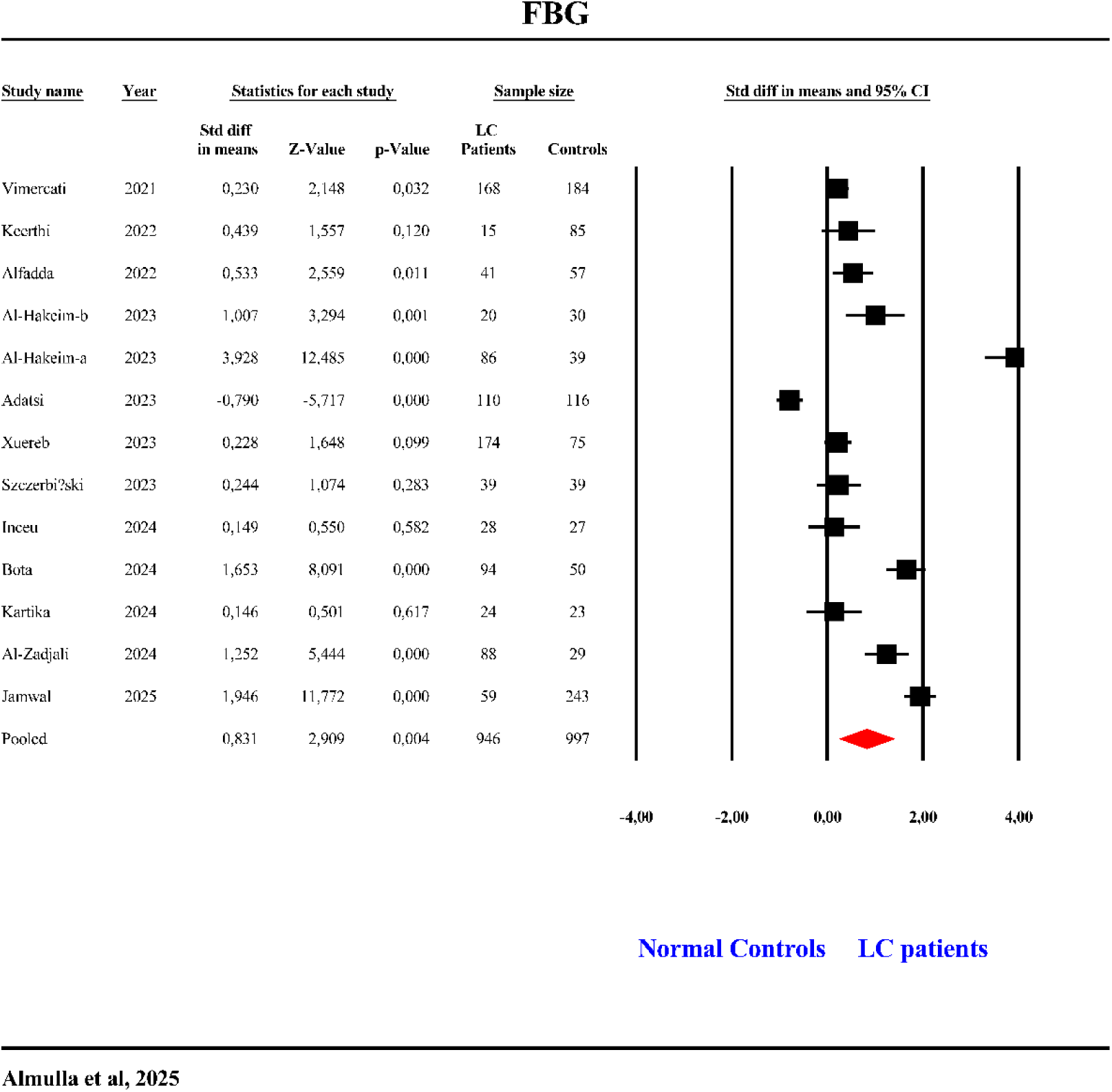
The forest plot of fasting blood glucose (FBG) in patients with Long COVID (LC) and normal controls.

**Figure 8.**
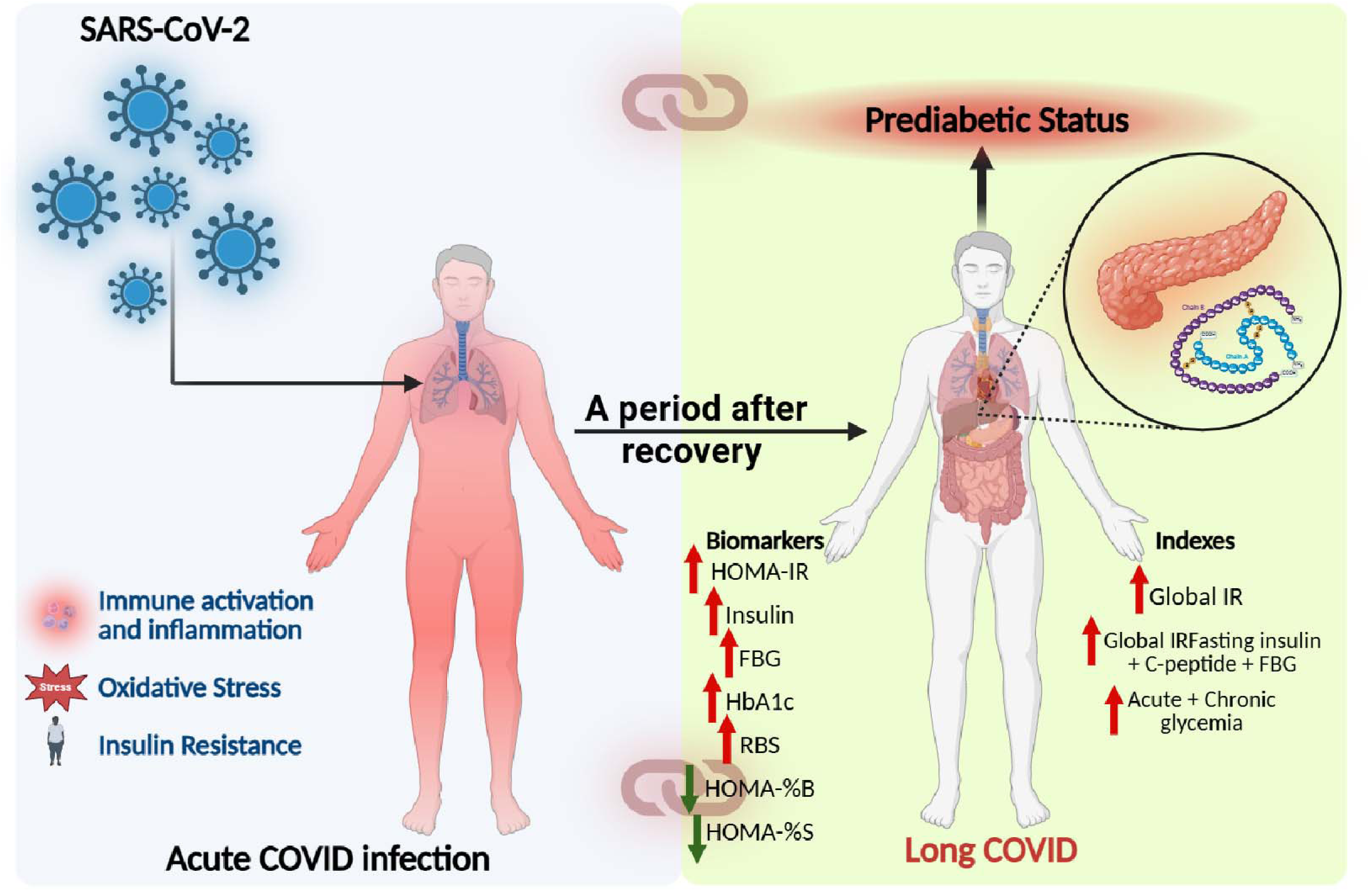
Summary of the insulin resistance (IR) in Long COVID (LC) disease. SARS-CoV-2: Severe acute respiratory syndrome coronavirus 2, HOMA-IR: Homeostatic Model Assessment for Insulin Resistance, FBG: fasting blood glucose, HbA1c: Hemoglobin A1C, RBS: Random blood sugar, HOMA-%B: Homeostasis Model Assessment for Beta-cell Function, HOMA-%S: Homeostatic Model Assessment for Insulin Sensitivity

## Secondary Outcomes Variables

### HbA1C

Nineteen studies investigated HbA1C (Table 1, ESF, Figure 1). One study showed CIs below zero, 8 above, and 10 overlapping (4 negative, 6 positive SMDs). LC patients demonstrated significantly higher HbA1C levels (Table 2, ESF, Figure 1), with no publication bias detected (Table 3).

### RBS

Thirty-six studies examined RBS (Table 1, ESF, Figure 2). Two studies had CIs below zero, 16 above, and 18 overlapping (9 negative, 9 positive SMDs). LC patients exhibited significantly higher RBS levels (Table 2). Egger’s and Kendall tests indicated no publication bias (Table 3).

### HOMA-%B

In this study, data from twenty-seven studies were combined to analyze HOMA-%B (Table 1, ESF, Figure 3). Seven studies reported CIs entirely below zero, one reported CIs entirely above zero, and nineteen showed overlapping CIs, with eight indicating negative and eleven indicating positive SMDs. Overall, LC patients demonstrated a significant reduction in HOMA-%B compared with normal controls (Table 2). Egger’s and Kendall tests confirmed the absence of publication bias (Table 3).

### HOMA-%S

Three studies assessed HOMA-%S (Table 1, ESF, Figure 4). All three studies showed CIs entirely below zero, indicating significantly reduced HOMA-%S in LC patients (Table 2). No publication bias was detected (Table 3).

### C-peptide

Four studies evaluated C-peptide levels (Table 1). Table 2 indicates no significant difference in C-peptide concentrations between LC patients and normal controls.

### Adiponectin

Four studies assessed adiponectin levels (Table 1). Table 2 showed no significant difference in adiponectin concentrations between LC patients and normal controls.

### Leptin

Five studies assessed leptin levels (Table 1). Table 2 revealed no significant difference in leptin concentrations between LC patients and normal controls.

### Ghrelin

Three studies evaluated ghrelin levels (Table 1). Table 2 indicated no significant difference in ghrelin concentrations between LC patients and normal controls.

## Meta-Regression Analysis

Meta-regression analysis explored sources of heterogeneity among studies investigating insulin resistance in LC (ESF, Table 4). Hospitalization during the acute illness significantly influenced most outcomes. Similarly, a post-acute period exceeding six months—the phase during which post-COVID manifestations emerge—had a marked effect. Other covariates, including medication use, age, ICU admission rate during acute illness, latitude, serum medium, sample size, and smoking status, also contributed to variability across outcomes.

## Discussion

### Persistent Insulin Resistance and Hyperglycemia in Long COVID

This meta-analysis is, to our knowledge, the first to comprehensively examine insulin resistance indices and related biomarkers in LC compared with normal controls. Our first major finding is that LC is associated with elevated IR across composite and solitary measures. Composite indices, including global IR, the fasting insulin + C-peptide + FBG, and the acute + chronic glycemia composite, were all increased in LC compared to normal controls. Solitary biomarkers showed convergent abnormalities, with higher HOMA-IR, insulin, HbA1c, FBG, RBS, alongside reduced HOMA-%S. Together, these results indicate a phenotype marked by hyperglycemia, compensatory upregulation of insulin secretion, and reduced tissue sensitivity to insulin.

The current findings are in line with prior evidence demonstrating increased IR in LC (Al-Hakeim, Al-Rubaye et al. 2023, Al-Hakeim, Khairi Abed et al. 2023, Maes, Almulla et al. 2023, Al Masoodi, Radhi et al. 2025, Fierro, Martín et al. 2025) and suggest that COVID-19 may increase diabetes risk by aggravating IR. COVID-19–related metabolic dysfunction might clinically manifest as new-onset diabetes, asymptomatic IR, and glucose intolerance (Montefusco, Ben Nasr et al. 2021). Man et al. demonstrated a markedly increased risk of IR and diabetes, reporting that up to 75% of patients with long COVID developed diabetes within one year of infection (Man, Andor et al. 2024).

Various mechanisms may contribute to IR and hyperglycemia in LC. Notably, LC-related IR was predicted by elevated peak body temperature during acute illness, suggesting the role of acute-phase inflammatory severity in subsequent metabolic outcomes including insulin resistance (Al-Hakeim, Al-Rubaye et al. 2023). In general, viral infections may induce IR; in murine models, IFN-γ might downregulate muscle insulin receptors, leading to compensatory hyperinsulinemia (Šestan, Marinović et al. 2018). Furthermore, in LC, instances of viral persistence have been reported (Vojdani, Vojdani et al. 2023, Vojdani, Almulla et al. 2024). SARS-CoV-2 invasion of pancreatic β-cells and other metabolic tissues can disrupt insulin production and signaling (Mahmudpour, Vahdat et al. 2022, Grubišić, Švitek et al. 2023). Virus-induced metabolic reprogramming towards anaerobic respiration, accompanied by acid-base disturbances, may exacerbate IR (van der Togt and Rossman 2023). SARS-CoV-2 infection of adipose tissue and β-cells promotes adipose dysfunction (e.g., reduced adiponectin) and β-cell impairment, both contributing to IR (Reiterer, Rajan et al. 2021). SARS-CoV-2 inhibits PGC-1α and irisin, leading to compromised mitochondrial function and energy metabolism, which results in decreased glucose uptake and increased IR (Mahmudpour, Vahdat et al. 2022, Khalil, Atia et al. 2025).

In LC, elevated cytokines may drive serine phosphorylation of IRS-1 which impairs insulin signaling and glucose uptake (Almulla, Thipakorn et al. 2024a, Berbudi, Khairani et al. 2025). Ongoing immune activation, including increased NETosis and altered microRNAs such as miR-21-5p, has been linked to post-COVID IR (Cabrera-Garcia, Quiroga et al. 2023). Sustained inflammatory, oxidative, and nitrosative stress signaling—such as activation of NF-κB and cytokines—can disrupt insulin receptor pathways (Keane, Cruzat et al. 2015, Onyango 2018). However, Al-Hakeim et al. reported that, in LC, inflammation during the acute phase exerts only a minor effect on new-onset IR, whereas activated IO&NS pathways during the chronic phase appear to have no significant impact (Al-Hakeim, Al-Rubaye et al. 2023). Upregulation of iNOS results in the S-nitrosylation and nitration of insulin signaling proteins (IR, IRS-1/2, Akt), which facilitates their inactivation and degradation (Kaneki, Shimizu et al. 2007).

LC is also marked by a pro-atherogenic lipid profile with increased TG, LDL, total cholesterol, and ApoB and decreased HDL and ApoA (Almulla, Thipakorn et al. 2025). HDL-C exhibits a positive correlation with insulin sensitivity, while reduced levels of HDL-C are linked to increased HOMA-IR (Goff, D’Agostino et al. 2005, Peng, Chiu et al. 2010). Furthermore, IR disrupts lipoprotein metabolism, resulting in elevated triglycerides and decreased HDL cholesterol, characteristic of the dyslipidemic profile associated with metabolic syndrome and T2DM (Bjornstad and Eckel 2018). Altered activity of hepatic and lipoprotein lipases in IR states further reduces HDL-C (de Vries, Borggreve et al. 2003).

LC is also accompanied by liver injury with elevated ALT and AST (Almulla, Thipakorn et al. 2024c), and IR associates with higher ALT and the AST/ALT ratio (Simental-Mendía, Rodríguez-Morán et al. 2017). Downregulation of Sirt1 in COVID-19 patients impairs insulin signaling and elevates oxidative stress (Wang, Kim et al. 2011, Izadpanah, Mudd et al. 2023). Gut dysbiosis following COVID-19 may further disturb metabolic regulation and aggravate IR (Bose, Bisht et al. 2025).

### β-Cell Dysfunction and Adipokine Levels in LC Disease

Our second major finding shows reduced HOMA-%B in LC, with no significant differences in adiponectin, ghrelin, or leptin. These results point to impaired β-cell function and a prediabetic state. Evidence supporting β-cell dysfunction in LC exists (Adatsi, Bockarie et al. 2023). However, the literature is mixed: some studies reported no significant change in HOMA-%B (Al-Hakeim, Al-Rubaye et al. 2023, Kartika, Subekti et al. 2024), whereas others observed increased HOMA-%B (Montefusco, Ben Nasr et al. 2021, Al Masoodi, Radhi et al. 2025). These discrepancies likely reflect heterogeneity in disease stage, acute-phase severity, treatment exposures, and recovery trajectories, but our pooled result favors β-cell impairment.

SARS-CoV-2 infects β-cells through ACE2, leading to direct cytopathic effects and reduced insulin secretion (Afrisham, Jadidi et al. 2023). LC exhibits significant immune activation characterized by elevated levels of cytokines, chemokines, and growth factors, which may intensify β-cell stress and contribute to cell loss (Eizirik, Colli et al. 2009, Almulla, Thipakorn et al. 2024a). Cytokines, including IFN-γ, have the capacity to inhibit insulin gene expression and β-cell functionality (Pondugala, Sasikala et al. 2015). Autoimmune mechanisms may also contribute; LC is associated with increased autoantibodies targeting self-proteins (Vojdani, Almulla et al. 2024, Almulla, Maes et al. 2024d), consistent with post-viral immune dysregulation that could involve islet antigens.

Molecular mimicry presents an intriguing mechanism for the rapid onset of islet autoimmunity. Viral epitopes that share structural characteristics with islet antigens may enhance pre-existing autoimmune tendencies, accelerating the onset of T1DM instead of initiating autoimmunity from the beginning (Christen, Edelmann et al. 2004). Extended β-cell infection may enhance MHC-I expression, thereby maintaining antigen presentation and facilitating epitope spreading as further islet antigens are revealed (Boddu, Aurangabadkar et al. 2020). This cascade promotes the activation of cytotoxic CD8+ T-cells and the diversification of autoantibody responses to insulin, GAD, and IA-2, leading to progressive β-cell loss and hyperglycemia (Taplin and Barker 2008). Increased NET formation during COVID-19 may exacerbate autoimmune inflammation and adjacent tissue damage, thereby compromising islet integrity (Vorobjeva and Chernyak 2020). All in all, these findings indicate a trajectory in which inflammation, oxidative stress, and potential autoimmunity intersect with pancreatic function.

### LC Disease Predisposes Patients to Diabetes Mellitus and Its Consequences

Epidemiologic data indicates increased risk after COVID-19 for new-onset diabetes (types 1 and 2), metabolic syndrome, and related complications (Zhang, Mei et al. 2022, Kim, Arora et al. 2023). Risk peaks in the first three months post-infection yet remains elevated over the longer term (Ssentongo, Zhang et al. 2022, Zhang, Mei et al. 2022). Research indicates that this risk may be partially independent of conventional factors like BMI, suggesting a direct impact from COVID-19 itself (Zhang, Mei et al. 2022, Grubišić, Švitek et al. 2023). Indeed, in our meta-regression analysis we didn’t find any impact for BMI on the outcomes. These observations are consistent with our aggregated evidence of persistent IR and β-cell dysfunction in LC, as well as the mechanistic connections to inflammation, oxidative stress, endothelial dysfunction, and possible autoimmunity outlined previously.

The clinical implications are evident. Individuals recovering from COVID-19, especially those with LC features, may require metabolic follow-up to detect emergent IR, dysglycemia, dyslipidemia, and related metabolic aberrations, including hepatic involvement (Almulla, Thipakorn et al. 2024c). The convergence of adverse lipid profiles, insulin signaling defects, and β-cell stress argues for integrated management strategies that address immune activation, oxidative pathways, and metabolic regulation simultaneously. While definitive interventional trials in LC remain limited, the mechanistic and epidemiologic signals justify proactive monitoring and targeted risk modification.

Finally, this integrated perspective underscores the necessity for multidisciplinary LC care pathways and encourages additional research to enhance phenotyping, identify modifiable targets, and evaluate interventions that influence neuro-immune – metabolic – oxidative (NIMETOX) pathways-related processes(Maes, Almulla et al. 2025).

## Limitations

Although most results of this meta-analysis were not affected by publication bias, several limitations should be acknowledged. First, the analysis was constrained by the limited data available in the included studies, which prevented assessment of the effects of treatments administered during the acute phase or post-recovery period and vaccination state. Future research should investigate how therapeutic interventions during these stages influence LC outcomes. Second, the study would have been strengthened by the inclusion of additional components of the immune-metabolic-oxidative stress pathways (Maes, Almulla et al. 2025). Future investigations should aim to examine multiple related biomarkers simultaneously to better characterize their contribution to the pathophysiology of LC.

Third, due to insufficient reporting, the influence of pre-existing diabetes mellitus or other metabolic disorders during the acute infection phase could not be evaluated. Subsequent studies should provide detailed information on comorbid conditions such as obesity, metabolic syndrome, and related factors to enable a more comprehensive understanding of their role in LC–associated insulin resistance.

## Conclusion

The current meta-analysis is the first comprehensive evidence demonstrating that LC disease is characterized by persistent IR, hyperglycemia, and β-cell dysfunction, reflecting sustained metabolic disruptions beyond the acute infection phase

## Data Availability

The corresponding author (MM) will evaluate reasonable requests for access to the dataset (Excel file) used in this meta analysis, after all contributing authors have completed their data usage.

## Ethical approval and consent to participate

Not applicable.

## Consent for publication

Not applicable.

## Availability of data and materials

The corresponding author (MM) will evaluate reasonable requests for access to the dataset (Excel file) used in this meta-analysis, after all contributing authors have completed their data usage.

## Author’s contributions

AA and MM conceptualized and designed the study. Data collection was conducted collaboratively by AA and YZ. Statistical analyses were performed by AA and MM. All authors contributed to drafting the manuscript, reviewed it critically for important intellectual content, and approved the final version for submission.

## Acknowledgments

Not applicable.

