## supplementary file for "Increased Insulin Resistance and Hyperglycemia in Long COVID disease: A Systematic Review and Meta-Analysis"

**Running Title:** Insulin Resistance and Hyperglycemia in Long COVID.

(1-2) Abbas F. Almulla, (1,2) Yingqian Zhang, (3,4) Chavit Tunvirachaisakul, (5) Andre F Carvalho, (1-3,6-8) Michael Maes*

1. Sichuan Provincial Center for Mental Health, Sichuan Provincial People's Hospital, School of Medicine, University of Electronic Science and Technology of China, Chengdu 610072, China

2. Key Laboratory of Psychosomatic Medicine, Chinese Academy of Medical Sciences, Chengdu 610072, China

3. Department of Psychiatry, Faculty of Medicine, Chulalongkorn University, Bangkok, Thailand.

4. Cognitive Impairment and Dementia Research Unit, Faculty of Medicine, Chulalongkorn University, Bangkok, Thailand.

5. Innovation in Mental and Physical Health and Clinical Treatment (IMPACT) Strategic Research Centre, School of Medicine, Barwon Health, Deakin University, Geelong, VIC, Australia

6. Department of Psychiatry, Medical University of Plovdiv, Plovdiv, Bulgaria.

7. Research Institute, Medical University Plovdiv, Plovdiv, Bulgaria.

8. Kyung Hee University, 26 Kyungheedae-ro, Dongdaemun-gu, Seoul 02447, Republic of Korea.

**Corresponding authors:**

Prof. Dr. Michael Maes, and Yingqian Zhang.

Sichuan Provincial Center for Mental Health

Sichuan Provincial People’s Hospital,

School of Medicine,

University of Electronic Science and Technology of China

Chengdu 610072

China

[Michael Maes Google Scholar profile](https://scholar.google.com/citations?user=1wzMZ7UAAAAJ&hl=en)

<https://scholar.google.co.th/citations?user=1wzMZ7UAAAAJ&hl=th&oi=ao>

Highly cited author: 2003-2023 (ISI, Clarivate)

ScholarGPS: Worldwide #1 in molecular neuroscience; #1/4 in pathophysiology

Expert worldwide medical expertise ranking, Expertscape (December 2022), worldwide:

#1 in CFS, #1 in oxidative stress, #1 in encephalomyelitis, #1 in nitrosative stress, #1 in nitrosation, #1 in tryptophan, #1 in aromatic amino acids, #1 in stress (physiological), #1 in neuroimmune; #2 in bacterial translocation; #3 in inflammation, #4-5: in depression, fatigue and psychiatry.

**ESF. Table 1.** Search sentences and terms were used in each database.

| **Database Name** | **Search Sentence** | **No. of Articles** |
| --- | --- | --- |
| **PubMed/Medline** | (("Leptin"[Mesh] OR "Adiponectin"[Mesh] OR "Resistin"[Mesh] OR "Visfatin"[Title/Abstract] OR "Ghrelin"[Mesh] OR "Insulin"[Mesh] OR "Insulin Resistance"[Mesh] OR "HOMA-IR"[Title/Abstract] OR "fasting insulin"[Title/Abstract]) AND ("Long COVID"[Title/Abstract] OR "post-COVID"[Title/Abstract] OR "post-acute COVID-19 syndrome"[Title/Abstract])) NOT (gene[Title/Abstract] OR genes[Title/Abstract] OR genetic*[Title/Abstract] OR polymorphism*[Title/Abstract] OR "systematic review"[Title/Abstract] OR "meta-analysis"[Title/Abstract] OR review[Publication Type]) AND ("2021/01/01"[Date - Publication] : "2025/12/31"[Date - Publication]) | **26** |
|  | (("C-Peptide"[Mesh] OR "Glucagon-Like Peptide 1"[Mesh] OR "glucose metabolism"[Title/Abstract] OR "adipokines"[Title/Abstract] OR "HOMA index"[Title/Abstract]) AND ("post-COVID-19 syndrome"[Title/Abstract] OR "PASC"[Title/Abstract] OR "post-acute sequelae of SARS-CoV-2"[Title/Abstract])) NOT (genetic*[Title/Abstract] OR polymorphism*[Title/Abstract] OR "systematic review"[Title/Abstract] OR "meta-analysis"[Title/Abstract] OR review[Publication Type]) AND ("2021/01/01"[Date - Publication] : "2025/12/31"[Date - Publication]) | **6** |
|  | (("Metabolic Biomarkers"[Title/Abstract] OR "Hormonal Biomarkers"[Title/Abstract] OR "insulin sensitivity"[Title/Abstract] OR "adipokines"[Title/Abstract] OR "leptin"[Title/Abstract] OR "insulin resistance marker"[Title/Abstract]) AND ("Long COVID"[Title/Abstract] OR "post COVID"[Title/Abstract] OR "PASC"[Title/Abstract])) NOT (genetic*[Title/Abstract] OR gene[Title/Abstract] OR polymorphism*[Title/Abstract] OR "meta-analysis"[Title/Abstract] OR review[Publication Type]) AND ("2021/01/01"[Date - Publication] : "2025/12/31"[Date - Publication]) | **11** |
| **Google Scholar** | "leptin" OR "adiponectin" OR "resistin" OR "visfatin" OR "ghrelin" OR "GLP-1" OR "insulin" OR "insulin resistance" OR "HOMA-IR" OR "fasting insulin") AND ("Long COVID" OR "post-COVID" OR "post-acute COVID-19") -gene -genetic* -polymorphism* -"systematic review" -"meta-analysis" -review | **720** |
|  | "glucagon-like peptide-1" OR "C-peptide" OR "HOMA index" OR "glucose metabolism" OR "fasting glucose" OR "adipokine" OR "hormonal biomarker") AND ("post-COVID-19 syndrome" OR "post-acute sequelae of SARS-CoV-2" OR "PASC") -gene -genes -genetic* -review -"meta-analysis" | **20** |
|  | "metabolic hormone" OR "insulin resistance marker" OR "adipokines" OR "leptin" OR "insulin sensitivity" OR "plasma insulin") AND ("Long COVID" OR "PASC" OR "post COVID") -genetic* -genes -polymorphism* -review -"systematic review" -"meta-analysis" | **78** |
| **SCOPUS** | TITLE-ABS-KEY(leptin OR adiponectin OR resistin OR visfatin OR ghrelin OR "GLP-1" OR "glucagon like peptide 1" OR insulin OR "insulin resistance" OR "HOMA IR" OR "HOMA index" OR "fasting insulin")  AND TITLE-ABS-KEY("Long COVID" OR "post COVID" OR "post COVID 19" OR "post acute sequelae of SARS CoV 2" OR PASC) AND PUBYEAR > 2020 AND PUBYEAR < 2026 AND NOT TITLE-ABS-KEY(gene OR genes OR genetic OR polymorphism OR "systematic review" OR "meta analysis" OR review) | **386** |
|  | TITLE-ABS-KEY(metabolic AND hormone OR adipokine OR glucose AND metabolism OR insulin AND sensitivity OR c-peptide OR homa AND index) AND TITLE-ABS-KEY("long covid" OR "post covid" OR "post covid syndrome" OR "post acute covid" OR pasc) AND PUBYEAR > 2020 AND PUBYEAR < 2026 AND  NOT TITLE-ABS-KEY(gene OR genes OR genetic OR polymorphism OR systematic AND review OR meta AND analysis OR review) | **6** |

**ESF. Table 2.** Immune cofounder’s scale (ICS) applied from Andrés-Rodríguez. et al.. 2019

| **Methodological quality of the study** | |
| --- | --- |
| **1** | Study sample ≥ 128 participants including patients and controls (1= Yes. 0 = No) |
| **2** | Did the study control the results for potential confounders (e.g.. age. BMI. gender. race)? (1= Yes. 0 = No) |
| **3** | Were participants with Long COVID and controls age- and-gender-matched or was there a statistical control? (1= Yes. 0 = No) |
| **4** | Was the time of sample collection specified (e.g.. morning vs. evening)? (1= Yes. 0 = No) |
| **5** | Were participants with Long COVID free of immunomodulatory drugs including anti-cytokines. glucocorticoids. immunoglobulins. and immunosuppressants. or was there a medication washout period. or was drug intake statistically controlled for? (1= Yes. 0 = No) |
| **6** | Were participants with Long COVID free of antidepressants and mood stabilizers or were the data statistically controlled for? (1= Yes. 0 = No) |
| **7** | Reporting either the manufacturer of the test or detection limit and coefficients of variation (1= Yes. 0 = No) |
| **8** | Reporting how data under detection limit were handled (1 = Yes. 0 = No) |
| **9** | Reporting % of the sample under detection limit (1=Yes. 0= No) |
| **10** | Reporting blood fraction (serum. plasma. culture supernatant or whole blood) (1= Yes. 0 = No) |
| **Total quality score (10 points)** | |
| **Biomarker confounders red points**  *The red points should not be given if the item is statistically controlled for* | |
| **1** | 3 red points for comorbid illnesses such as autoimmune disorders & other immune disorders including rheumatoid arthritis. psoriasis. inflammatory bowel disease. chronic obstructive pulmonary disease. multiple sclerosis |
| **2** | 3 red points for use of recreational drugs such as methamphetamine or opioids |
| **3** | 2 red points when groups were not matched for age |
| **4** | 2 red points when groups were not matched for sex |
| **5** | 2 red points for medication use as for example immunomodulators |
| **6** | 2 red points for early traumatic life events |
| **7** | 2 red points for shift work and primary sleep disorders |
| **8** | 1.5 red points for use of antipsychotics |
| **9** | 1 red point for more common systemic immune disorders including diabetes type 1/2. essential hypertension. metabolic syndrome |
| **10** | 1 red point for not fasting (8 hours before blood extraction) |
| **11** | 1 red point for use of omega-3 and antioxidant supplements |
| **12** | 1 red point when data were not controlled for body mass index |
| **13** | 1 red point when data were not controlled for physical activity or sedentary life |
| **14** | 1 red point when data were not controlled for smoking |
| **15** | 1 red point for use of oral contraceptives or NSAIDs |
| **16** | 0.5 red points when data were not controlled for ethnicity in countries such as US. Brazil |
| **17** | 0.5 red points when data were not controlled for seasonality |
| **18** | 0.5 red points when data were not controlled for diurnal variation (8-10 a.m. versus all other time points) |
|  | **Total red point score (26 points)** |

**ESF. Table 3.** PRISMA checklist

| **Section/topic** | **#** | **Checklist item** | **Reported on page #** |
| --- | --- | --- | --- |
| **TITLE** | | | |
| Title | 1 | Identify the report as a systematic review. meta-analysis. or both. |  |
| **ABSTRACT** | | | |
| Structured summary | 2 | Provide a structured summary including. as applicable: background; objectives; data sources; study eligibility criteria. participants. and interventions; study appraisal and synthesis methods; results; limitations; conclusions and implications of key findings; systematic review registration number. |  |
| **INTRODUCTION** | | | |
| Rationale | 3 | Describe the rationale for the review in the context of what is already known. |  |
| Objectives | 4 | Provide an explicit statement of questions being addressed with reference to participants. interventions. comparisons. outcomes. and study design (PICOS). |  |
| **METHODS** | | | |
| Protocol and registration | 5 | Indicate if a review protocol exists. if and where it can be accessed (e.g.. Web address). and. if available. provide registration information including registration number. |  |
| Eligibility criteria | 6 | Specify study characteristics (e.g.. PICOS. length of follow-up) and report characteristics (e.g.. years considered. language. publication status) used as criteria for eligibility. giving rationale. |  |
| Information sources | 7 | Describe all information sources (e.g.. databases with dates of coverage. contact with study authors to identify additional studies) in the search and date last searched. |  |
| Search | 8 | Present full electronic search strategy for at least one database. including any limits used. such that it could be repeated. |  |
| Study selection | 9 | State the process for selecting studies (i.e.. screening. eligibility. included in systematic review. and. if applicable. included in the meta-analysis). |  |
| Data collection process | 10 | Describe method of data extraction from reports (e.g.. piloted forms. independently. in duplicate) and any processes for obtaining and confirming data from investigators. |  |
| Data items | 11 | List and define all variables for which data were sought (e.g.. PICOS. funding sources) and any assumptions and simplifications made. |  |
| Risk of bias in individual studies | 12 | Describe methods used for assessing risk of bias of individual studies (including specification of whether this was done at the study or outcome level). and how this information is to be used in any data synthesis. |  |
| Summary measures | 13 | State the principal summary measures (e.g.. risk ratio. difference in means). |  |
| Synthesis of results | 14 | Describe the methods of handling data and combining results of studies. if done. including measures of consistency (e.g.. I^2^) for each meta-analysis. |  |
| Risk of bias across studies | 15 | Specify any assessment of risk of bias that may affect the cumulative evidence (e.g.. publication bias. selective reporting within studies). |  |
| Additional analyses | 16 | Describe methods of additional analyses (e.g.. sensitivity or subgroup analyses. meta-regression). if done. indicating which were pre-specified. |  |
| **RESULTS** | | |  |
| Study selection | 17 | Give numbers of studies screened. assessed for eligibility. and included in the review. with reasons for exclusions at each stage. ideally with a flow diagram. |  |
| Study characteristics | 18 | For each study. present characteristics for which data were extracted (e.g.. study size. PICOS. follow-up period) and provide the citations. |  |
| Risk of bias within studies | 19 | Present data on risk of bias of each study and. if available. any outcome level assessment (see item 12). |  |
| Results of individual studies | 20 | For all outcomes considered (benefits or harms). present. for each study: (a) simple summary data for each intervention group (b) effect estimates and confidence intervals. ideally with a forest plot. |  |
| Synthesis of results | 21 | Present results of each meta-analysis done. including confidence intervals and measures of consistency. |  |
| Risk of bias across studies | 22 | Present results of any assessment of risk of bias across studies (see Item 15). |  |
| Additional analysis | 23 | Give results of additional analyses. if done (e.g.. sensitivity or subgroup analyses. meta-regression [see Item 16]). |  |
| **DISCUSSION** | | |  |
| Summary of evidence | 24 | Summarize the main findings including the strength of evidence for each main outcome; consider their relevance to key groups (e.g.. healthcare providers. users. and policy makers). |  |
| Limitations | 25 | Discuss limitations at study and outcome level (e.g.. risk of bias). and at review-level (e.g.. incomplete retrieval of identified research. reporting bias). |  |
| Conclusions | 26 | Provide a general interpretation of the results in the context of other evidence. and implications for future research. |  |
| **FUNDING** | | |  |
| Funding | 27 | Describe sources of funding for the systematic review and other support (e.g.. supply of data); role of funders for the systematic review. |  |

**ESF. table 4.** Characteristics of the studies included in the systematic reviews and meta-analysis.

| **NO** | **Authors. years** | **Setting** | **Post COVID period-Months** | **Type of case** | **Type of Control** | **Sample Size** | | | **Age** | | **Specimen** | **Quality score** | **Red point score** | **Findings** |
| --- | --- | --- | --- | --- | --- | --- | --- | --- | --- | --- | --- | --- | --- | --- |
|  |  |  |  |  |  | **Cases M/F** | **Control M/F** | **Total M/F** | **Case-Mean (SD)** | **Control- Mean (SD)** |  |  |  |  |
| 1 | (Holmes, Wist et al. 2021) | Australia | Approximately 3 months | Post-acute phase nonhospitalized COVID-19 patients. | Healthy controls | NA | NA | NA | NA | NA | Plasma | 4 | 14.5 | Glucose* |
| 2 | (Silva, Pereira et al. 2023) | Brazil | 70.50 ± 43.10 days post-diagnosis | Mild-to-moderate post-COVID-19 patients | Healthy age-matched controls, tested negative for SARS-CoV-2 | 20 11/9 | 20 14/6 | 40 25/15 | 29.41 (21.90– 34.96) | 29.39 (21.25 – 32.62) | Serum | 5 | 4.5 | Adiponectin#, Glucsoe#, HOMA-IR*, Insulin*, Leptin#, PAI-1#, TG# |
| 3 | (Oliván-Blázquez, Bona-Otal et al. 2024) | Spain | 12–24 months | Patients with post-COVID condition | Individuals recovered within three months from acute COVID-19 | 85 17/68 | 85 66/19 | 170 83/87 | 47 (10) | 48 (10) | Plasma | 7 | 12 | HbA1c*, TG# |
| 4 | (Agafonova, Elovikova et al. 2024) | Russia | 12 months for the first stage | Patients with a history of COVID-19 confirmed by positive PCR | Subjects with a negative PCR test for COVID-18 | 138 27/111 | 87 18/69 | 225 45/180 | 61 (47-70) | 59 (44-67) | Blood | 7 | 6 | Glucose# |
| 5 | (Alfadda, Rafiullah et al. 2022) | Saudi Arabia | 6 months | With at least one symptom | No symptoms | NA | NA | NA | 51.34 (18,2) | 46.44 (16,8) | Serum | 3.5 | 13.5 | FBG#, HbA1c#, TG#, TG_1#, Vitamin D#, Vitamin D_1# |
| 6 | (Alshehri, AlQahtani et al. 2023) | Saudi Arabia | 1 month | Residual Neurological Deficits | Complete Recovery | 12 9/3 | 13 6/7 | 25 15/10 | NA | NA | NA | 3 | 19 | HbA1c*, TG* |
| 7 | (Abdulaziz Alsufyani 2023) | Saudi Arabia | 6‐month | after acute infection | non COVID infected patients | 37 37/0 | 35 35/0 | 72 72/0 | 11 (1202) | 10.86 (1089) | Serum | 7 | 6 | Glucose#, TG# |
| 8 | (Al-Zadjali, Al-Lawati et al. 2024) | Oman | 3-6 months | Long-COVID-19 patients-Mild-Moderate | Healthy individuals who had neither been affected by COVID-19 nor vaccinated. | 88 52/36 | 29 13/16 | 117 65/52 | 39.66 (10.95) | 38.21 (10.55) | Serum | 4 | 14.5 | FBG#, FBG_1#, HbA1c#, HbA1c_1#, HOMA-IR#, HOMA-IR_1#, Insulin#, Insulin_1#, oxLDL*, oxLDL_1# |
| 9 | (Duran, Kurtipek et al. 2022) | Turkey | At least one year after hospitalization. | Patients diagnosed with long COVID | Healthy individuals with a prior history of COVID-19 who fully recovered | 52 26/26 | 80 28/52 | 132 54/78 | 53.6 (12.8) | 49.0 (12.3) | Serum | 2 | 19.5 | Glucose#, TG* |
| 10 | (Emiroglu, Dicle et al. 2024) | Turkey | 0-11 months | Post-COVID-19 syndrome patients with hyperglycemia or diabetes. | Post-COVID-19 syndrome patients with normal fasting blood glucose. | 103 34/69 | 408 208/200 | 511 242/269 | 57.86 (10.33) | 50.03 (13.24) | Serum | 2.5 | 25 | HbA1c#, TG# |
| 11 | (Erol, Tezcan et al. 2023) | Turkey | At least one year after laboratory-confirmed COVID-19. | Long COVID patients with cardiac symptoms. | Age- and gender-matched individuals without a history of COVID-19. | 105 45/60 | 184 83/101 | 289 128/161 | 56.1 (11.3) | 55.8 (10.7) | Blood | 4 | 6.5 | Glucose#, TG# |
| 12 | (Korkmaz, Çınar et al. 2024) | Turkey | 3 to 6 months | Post-COVID-19 patients without hospitalization | Post-COVID-19 patients without hospitalization | 201 89/112 | 195 104/91 | 396 193/203 | 48.2 (16.3) | 49.2 (14.9) | blood | 4 | 13.5 | Glucose#, TG# |
| 13 | (Kuryłowicz, Babicki et al. 2024) | Poland | 12 and 16 weeks after the COVID‐19 | Post-COVID syndrome patients.IST | Post-COVID syndrome (PCS) patients.IST | 69 21/48 | 1280 491/789 | 1349 512/837 | 45.8 (11.6) | 51.6 (13.1) | Serum | 3 | 26 | Glucose#, TG# |
| 14 | (Labarca, Henríquez-Beltrán et al. 2022) | Chile | 4 months and 1 year | Obstructive Sleep Apnea | Non OSA | 33 22/11 | 23 10/13 | 56 32/24 | 51.4 (11.1) | 38.3 (12.1) | Serum | 4 | 19 | Glucose*, HOMA-IR#, TG# |
| 15 | (Mora, Kogut et al. 2023) | United States | Symptoms exceeding 28 days were classified as long COVID | Long COVID | No COVID | 94 28/66 | 104 49/55 | 198 77/121 | 43.8 (9.9) | 44.8 (12.3) | Blood | 5 | 21 | Glucose*, Glucose_1*, HbA1c#, HbA1c_1#, TG# |
| 16 | (Paris, Palomba et al. 2023) | Italy | Within 2 months | Post-COVID-19 patients with long-COVID conditions | Age- and sex-matched healthy volunteers | 38 35/3 | 38 35/3 | 76 70/6 | 58.82 (10.08) | 57.93 (11.23) | NA | 4 | 17 | Glucose#, Glucose_1#, Glucose_2#, Glucose_3#, TG#, TG_1# |
| 17 | (Zerón-Rugerio, Zaragozá et al. 2024) | Spain | NA | Patients with post-COVID ME-CFS | Matched healthy controls | 23 10/13 | 31 10/21 | 54 20/34 | 49.61 (2.09) | 43.06 (1.99) | Plasma | 4.5 | 9 | Glucose#, TG# |
| 18 | (Sumbalová, Kucharská et al. 2022) | Slovakia | NA | Post COVID | Healthy Control | 14 8/6 | 15 6/9 | 29 14/15 | 51.3 (2.3) | 51.3 (2.3) | Plasma | 4.5 | 14 | Glucose#, Glucose_1#, TG#, TG_1#, TG_2#, TG_3# |
| 19 | (Szczerbiński, Okruszko et al. 2023) | Poland | Approximately six months | Adults with a history of SARS-CoV-2 infection confirmed by PCR and hospitalized during the acute phase. | Age ,sex and BMI-matched participants from a population study conducted pre-pandemic. | 39 13/26 | 39 13/26 | 78 26/52 | 48.64 (2.24) | 48.79 (2.22) | Serum | 6 | 9 | CHO*, FBG#, HbA1c#, HOMA-2#, HOMA-B#, Insulin#, TG# |
| 20 | ((Tong, Yan et al. 2022), Yan et al. 2022) | China | 1 year (375.0 ± 11.0 days) | COVID-19 survivors one year after discharge | Age- and gender-matched healthy medical staff | 54 19/35 | 119 46/73 | 173 65/108 | 54 (41-61) | 52 (42-61) | Serum | 6 | 12 | Glucose#, Glucose_1#, TG*, TG_1* |
| 21 | (Tudoran, Bende et al. 2023) | Romania | Median time elapsed since COVID-19 diagnosis was 56 days for group A and 63 days for group B. | Group A: 67 Women with MS and a History of COVID-19 | Healthy, premenopausal, age-matched women who never had COVID-19. | 67 0/67 | 40 0/40 | 107 0/107 | 50.59 (4.53) | 49.47 (5.14) | Serum | 4.5 | 14.5 | Glucose#, Glucose_1*, TG#, TG_1#, TyG index#, TyG index_1# |
| 22 | (Verma, Ramayya et al. 2022) | United States | Mean 332 ± 130 days | Patients with post-COVID-19 syndrome PASC-CVS | Healthy controls | 23 2/21 | 23 10/13 | 46 12/34 | 46 (11) | 56 (12) | Serum | 4 | 6.5 | Glucose*, HbA1c*, TG# |
| 23 | (Vyas, Joshi et al. 2023) | India | 1 year | Post COVID-Hypertensive | postCOVID-Normotensive | 80 60/20 | 168 109/59 | 248 169/79 | 52.39 (12.64) | 50.35 (13.91) | Blood | 4 | 16.5 | HbA1c#, TG# |
| 24 | (Xuereb, Borg et al. 2023) | Malta | Approximately 6 months (median follow-up: 173.5 days). | Patients previously diagnosed with COVID-19 infection. | Age- and gender-matched individuals who tested negative for COVID-19. | 174 69/105 | 75 34/41 | 249 103/146 | 45.5 (35-58.75) | 44 (37.5-56.5) | Serum | 5.5 | 12.5 | FBG#, HbA1c#, Insulin#, TG# |
| 25 | (Yamamoto, Otsuka et al. 2023) | Japan | at least 6 months | Long COVID with ME-CSF | No fatigue | 50 24/26 | 95 38/57 | 145 62/83 | 42 (30.3–51.8) | 43 (29.5-51) | Blood | 4 | 18 | Glucose* |
| 26 | (Gupta, Nicholas et al. 2024) | UK | 7 | PostCOVID | Healthy Control | 21 15/6 | 10 7/3 | 31 22/9 | 54.28 (3.17) | 58 (52–67) | Serum | 4.5 | 13 | HbA1c# |
| 27 | (Meisinger, Goßlau et al. 2022) | Germany | 9.3 | Post COVID | Controls | 72 0/72 | 80 0/80 | 152 0/152 | 45.41 (16.26) | 45.11 (19.25) | blood | 8.5 | 11 | Glucose*, Glucose_1# |
| 28 | (Torki, Hoseininasab et al. 2024) | Iran | At least 3 | Post COVID | Controls | 88 43/45 | 96 55/41 | 184 98/86 | 47.45 (3.31) | 43 (37.25–58) | Blood | 4.5 | 9 | Glucose# |
| 29 | (Al Masoodi, Radhi et al. 2023) | Iraq | NA | Post COVID | Controls | 60 27/33 | 30 8/22 | 90 35/55 | 35.97 (9.17) | 33.23 (6.11) | Serum | 8 | 7.5 | Glucose#, HOMA-2#, HOMA-S*, Insulin# |
| 30 | (Clemente, Sinatti et al. 2022) | Italy | 3 | Post COVID | Controls | 48 34/14 | 45 22/23 | 93 56/37 | 62.54 (10.18) | 60.62 (17.37) | Serum | 3.5 | 19.5 | Glucose# |
| 31 | (Montefusco, Ben Nasr et al. 2021) | Italy | 2 months | Post-COVID | Healthy control | 10 7/3 | 15 10/5 | 25 17/8 | 46,9 (3.8) | 47.2 (3.1) | Serum | 5 | 11 | C-peptide#, HOMA-B#, HOMA-IR#, Insulin# |
| 32 | (Domingo et al., 2024) | Spain | More than 12 months | Long COVID | Healthy controls | 23 8/15 | 31 9/22 | 54 17/37 | 48.7 (2.4) | 41.7 (1.8) | Plasma | 9.5 | 7 | Adiponectin*, Leptin#, Serpin# |
| 33 | (Орлова, Ломайчиков et al. 2021) | Russia | NA | Patients with acute coronary syndrome who had previously suffered from COVID-19 | Patients with ACS without a history of COVID-19 | 109 109/0 | 76 76/0 | 185 185/0 | 64.4 (62, 0; 66, 9) | 68.2 (66, 2; 71, 4) | Serum | 2 | 20 |  |
| 34 | (Fernandez-de-las-Peñas, Notarte et al. 2024) | Spain | 6 | Post COVID | Controls | 300 140/160 | 112 73/39 | 412 213/199 | 63 (15) | 59.5 (17) | Serum | 5.5 | 10.5 | Glucose*, Glucose_1*, Glucose_2#, Glucose_3* |
| 35 | (Di Ciaula et al., 2024) | Italy | 12 months | Long COVID patients | Asymptomatic post-COVID controls | 265 128/137 | 120 68/52 | 385 196/189 | 50.5 (17) | 50 (14) | Serum | 9.5 | 3.5 | Leptin#, Resistin# |
| 36 | Al-Hakeim. Khairi Abed et al. 2023 | Iraq | 3-6 months | Long COVID (2groups) | Healthy Control | 20 12/8 | 30 16/14 | 70 34/36 | 32.5 (8.4) | 29.2 (8.1) | Serum | 7 | 9,5 | FBG#, HOMA-2#, HOMA-B#, HOMA-S*, Insulin# |
| 37 | (Al-Hakeim, Al-Rubaye et al. 2022b) | Iraq | 3-6 months | Long COVID | Healthy control | 86 62/24 | 39 24/15 | 125 86/39 | 28.4 (6.2) | 28.1 (7.6) | Serum | 7 | 7,5 | FBG#, Insulin# |
| 38 | (Atieh, Durieux et al. 2024) | USA | 29 months | COVID-19 survivors | No COVID-19 | 80 53/27 | 80 55/25 | 160 108/52 | 42.9  (13.6) | 43.2  (15.4) | Serum | 4 | 14.5 | HOMA-IR#, oxLDL# |
| 39 | (Bielecka-Dabrowa, Kapusta et al. 2024) | USA | 3-6 months | Long COVID females | Healthy females | 1227 0/1227 | 719 0/719 | 1946 0/1946 | 53  (14.84) | 52.14 (16.34) |  | 3 | 16 | Glucose#, TG#, Vitamin D* |
| 40 | (Jamwal, Chhabra et al. 2025) | USA | NA | Post-COVID with new-onset diabetes (NODAC) | COVID-19 with normal glucose | 59 31/28 | 243 142/101 | 302 173/129 | 54.71 (12.85) | 50.07 (17.17) | Serum | 4 | 19 | C-peptide*, FBG#, Glucose#, HbA1c#, HOMA-B*, HOMA-IR#, Insulin# |
| 41 | (Matviichuk, Yerokhovych et al. 2024) | USA | 6 months | T2DM with PCS | T2DM without PCS | 44 24/20 | 21 11/10 | 65 35/30 | 61.86 (11.32) | 62  (9.69) |  | 3 | 14.5 | HbA1c# |
| 42 | (Rajamanickam, Venkataraman et al. 2023) | USA | 3 months | Children post-SARS-CoV-2 | Healthy children | 19 11/8 | 20 14/6 | 39 25/14 |  |  | Plasma | 5 | 9 | Ghrelin#, Glucose#, PAI-1#, Resistin*, Visfatin* |
| 43 | (Jud, Gressenberger et al. 2021) | USA | 28,6 ± 3,0 weeks | Post-COVID | Controls | 14 7/7 | 14 7/7 | 28 14/14 | 68.7  (12) | 30.7  (4.2) | Serum | 3 | 17 | HbA1c# |
| 44 | (Bota, Bratosin et al. 2024) | USA | 6 months | Long COVID <65 y | No Long COVID | 71 37/34 | 50 27/23 | 121 64/57 |  |  | Serum | 3 | 15 | FBG#, FBG_1# |
| 45 | (Parás-Bravo, Fernández-de-Las-Peñas et al. 2024) | USA | 4-20 months | Cognitive loss | No cognitive loss | 0 / | 0 / | 0 0/0 | 69.0  (10.5) |  |  | 0 | 18 | Glucose*, Glucose_1*, Glucose_2* |
| 46 | (Ach, Ben Haj Slama et al. 2024) | USA | 11.5 months | Long COVID | Recovered post-COVID | 32 9/23 | 32 14/18 | 64 23/41 | 42.56 (13.45) | 43.31 (14.30) | Serum | 3 | 14 | Glucose* |
| 47 | (Adatsi, Bockarie et al. 2023) | USA | 6-8 months | Recovered COVID | No COVID | 110 45/65 | 116 61/55 | 226 106/120 | 37.06 (14.11) | 35.43 (11.84) | Serum | 3 | 18 | FBG*, HOMA-B*, HOMA-IR#, Insulin*, TyG index* |
| 48 | (Al Masoodi, Radhi et al. 2025) | USA | 17.3 ± 6.1 months | Long COVID | No Long COVID | 60 27/33 | 30 10/20 | 90 37/53 | 37.0  (9.4) | 35.8  (7.5) | Serum | 5 | 7 | Glucose#, HOMA-2#, HOMA-B#, HOMA-S*, Insulin# |
| 49 | (Barichello, Kluwe-Schiavon et al. 2025) | USA | 4-6 weeks | Post-COVID | Healthy controls | 25 9/16 | 57 10/47 | 82 63/63 | 38.04 (14.05) | 38.95 (13.41) | Plasma | 5 | 13 | Adiponectin#, Leptin#, Resistin* |
| 50 | (Duarte, Sambra et al. 2024) | USA | 2 months | Post-COVID NOD | Post-COVID no diabetes | 16 16/0 | 25 25/0 | 41 0/0 | 44.82 (42.26) | 38.00 (31.44) | Serum | 2 | 16 | C-peptide#, Glucose#, HbA1c#, Insulin* |
| 51 | (Ghosh, Niesen et al. 2024) | USA | 28–42 days | Long COVID | COVID+ no Long COVID | 36 36/0 | 17 17/0 | 53 0/0 |  |  | Serum | 4 | 16 | Glucose#, TG# |
| 52 | (López-Hernández, Oropeza-Valdez et al. 2023) | USA | 2 years | Recovered COVID (2020) | RT-qPCR negative | 20 11/9 | 15 9/6 | 35 15/15 | 51.8  (11.6) | 47.2  (8.4) | Plasma | 3 | 18 | Glucose#, Glucose_1*, HOMA-IR#, Insulin#, TG# |
| 53 | (Keerthi, Sushmita et al. 2022) | USA | 3 months | NODM post-COVID | No diabetes onset | 15 7/8 | 85 60/25 | 100 33/33 | 48.31 (19.07) | 48.31 (19.07) |  | 0 | 16 | C-peptide#, FBG#, HbA1c#, HOMA-B#, HOMA-IR#, Insulin# |
| 54 | (Matviichuk, Yerokhovych et al. 2024) | USA | 6 months | Post-COVID T2DM | T2DM without PCS | 227 103/124 | 242 127/115 | 469 239/239 | 60.64  (9.69) | 60.70 (10.44) | Blood | 0 | 15 | HbA1c# |
| 55 | (Struttmann, Shah et al. 2024) | USA | 18–663 days | Post-COVID ±PASC | Non-PASC post-COVID | 74 20/54 | 25 16/9 | 99 63/63 | 52.1  (13.7) |  | Serum | 3 | 21 | Glucose*, HbA1c* |
| 56 | (Vimercati, De Maria et al. 2021) | Italy | 35 days | HCWs post-COVID | HCWs no Long COVID | 168 71/97 | 184 77/107 | 352 204/204 | 45.2  (13.3) | 44.2  (13.2) |  | 0 | 20 | FBG#, TG# |
| 57 | (Kuryata, Mytrokhina et al. 2024) | Ukraine | 3-12 months | Post-COVID + IHD | Post-COVID + IHD (std therapy) | 31 17/14 | 24 9/15 | 55 29/29 | 46.08 (17.87) | 47.74 (16.15) |  | 1 | 16 | Glucose*, HOMA-IR*, IGF-1#, Insulin* |
| 58 | (Kartika, Subekti et al. 2024) | Indonesia | 12 months | COVID-19 survivors (12 mo) | Post-COVID normal HOMA-IR | 24 16/8 | 23 15/8 | 47 16/16 | 47.46 (12.39) | 47.78 (10.69) |  | 2 | 16.5 | TG#, TG_1# |
| 59 | (Manuilov and Mykhailovska 2024) | Ukraine | >3 months | Post-COVID + IHD | Non-COVID IHD | 31 31/0 | 15 15/0 | 46 0/0 | 69.71  (9.32) |  | Serum | 3 | 16.25 | Ghrelin*, Glucose#, Insulin#, Vasopressin# |
| 60 | (Visconti, Rocha et al. 2025) | Brazil | ≥4 months | Post-COVID (≥4 mo) | Post-COVID no fatigue | 57 36/21 | 58 27/31 | 115 52/52 |  |  | Serum | 2 | 20.5 | Adiponectin#, Irisin#, Leptin* |
| 61 | (Mittal, Ghosh et al. 2021) | India | 92 days | T2DM + COVID history | T2DM no COVID | 20 8/12 | 56 35/21 | 76 33/33 | 56.14 (11.37) | 58.33  (9.85) | Blood | 0 | 15.75 | HbA1c*, HbA1c_1# |
| 62 | (Inceu, Nechifor et al. 2024) | Romania | 1–27 months | Adults post-COVID | Healthy adults (pre-pandemic) | 28 10/18 | 27 8/19 | 55 37/37 | 27.64  (2.34) | 30.25  (5.87) | Plasma | 4 | 10.75 | FBG#, Ghrelin# |
| 63 | (Vojdani, Almulla et al. 2024) | Iraq | 3-6 months | Long COVID | Healthy adults (pre-pandemic) | 90 40/50 | 90 52/38 | 180 88/88 | 34.67 (14.53) | 38.24 (13.02) | Serum | 8 | 5.75 | HOMA-IR# |

*: Indicates that patients have reduced level of the measured metabolite compared to healthy control

^#:^ Indicates that patients have increased level of the measured metabolites compared to healthy control

**ESF, Table 4.** Results of Meta-regression

| Variables | No. of Studies | Covariates | 1-sided p-value | Z-Value |
| --- | --- | --- | --- | --- |
| Fasting insulin + C-peptide + fasting glucose/FBG | 11 | Medication-Free | 0.026 | 1.49 |
| Acute + Chronic glycemia | 50 | More than 6 Months | 0.0015 | -2.96 |
|  | 17 | Hospitalization during acute stage of illness | 0.0002 | 3.62 |
| Global IR | 11 | Hospitalization during acute stage of illness | 0.0002 | 3.52 |
|  | 53 | More than 6 months | 0.026 | -1.93 |
| HbA1c | 18 | More than 6 Months | 0.021 | -2.02 |
|  | 6 | Hospitalization during acute stage of illness | 0.0005 | 3.27 |
|  | 4 | Number of patients admitted to ICU | 0.0000 | 6.92 |
|  | 7 | Medication free-No | 0.008 | 2.41 |
| FBG | 7 | Medication free-all | 0.019 | 2.06 |
| Glucose | 35 | Latitude | 0.0094 | -2.35 |
|  | 30 | Less than 3 months | 0.020 | 2.05 |
|  | 11 | Hospitalization during acute stage of illness | 0.022 | 2.01 |
|  | 26 | Serum | 0.032 | -1.85 |
| HOMA-IR | 15 | Sample size | 0.0067 | 2.47 |
| HOMA-%B | 23 | More than 6 Months | 0.022 | 2.01 |
|  | 8 | Smoking% | 0.011 | 2.28 |

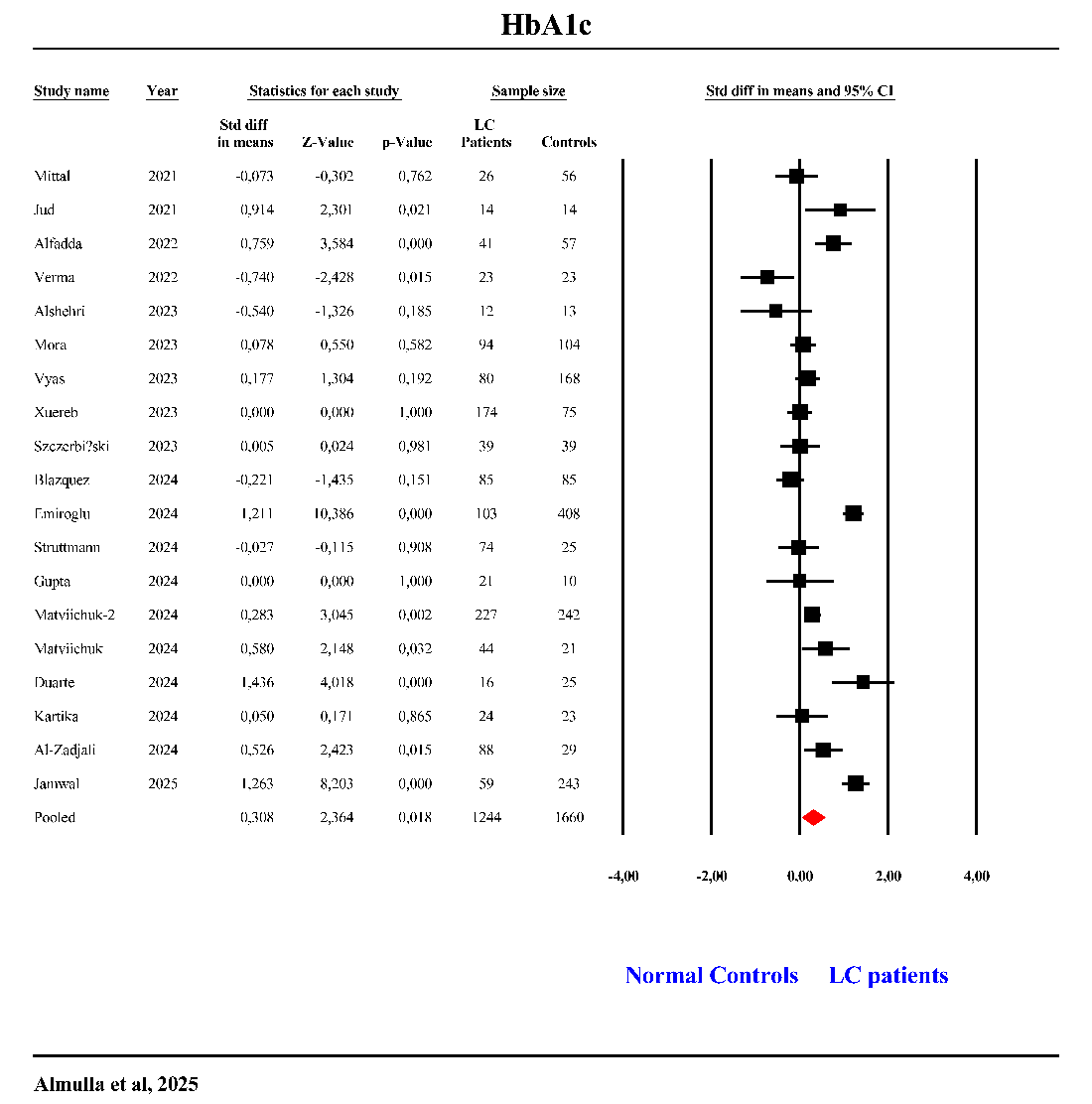

ESF, Figure 1. The forest plot of glycated hemoglobin (HbA1c) in patients with Long COVID (LC) and normal controls.

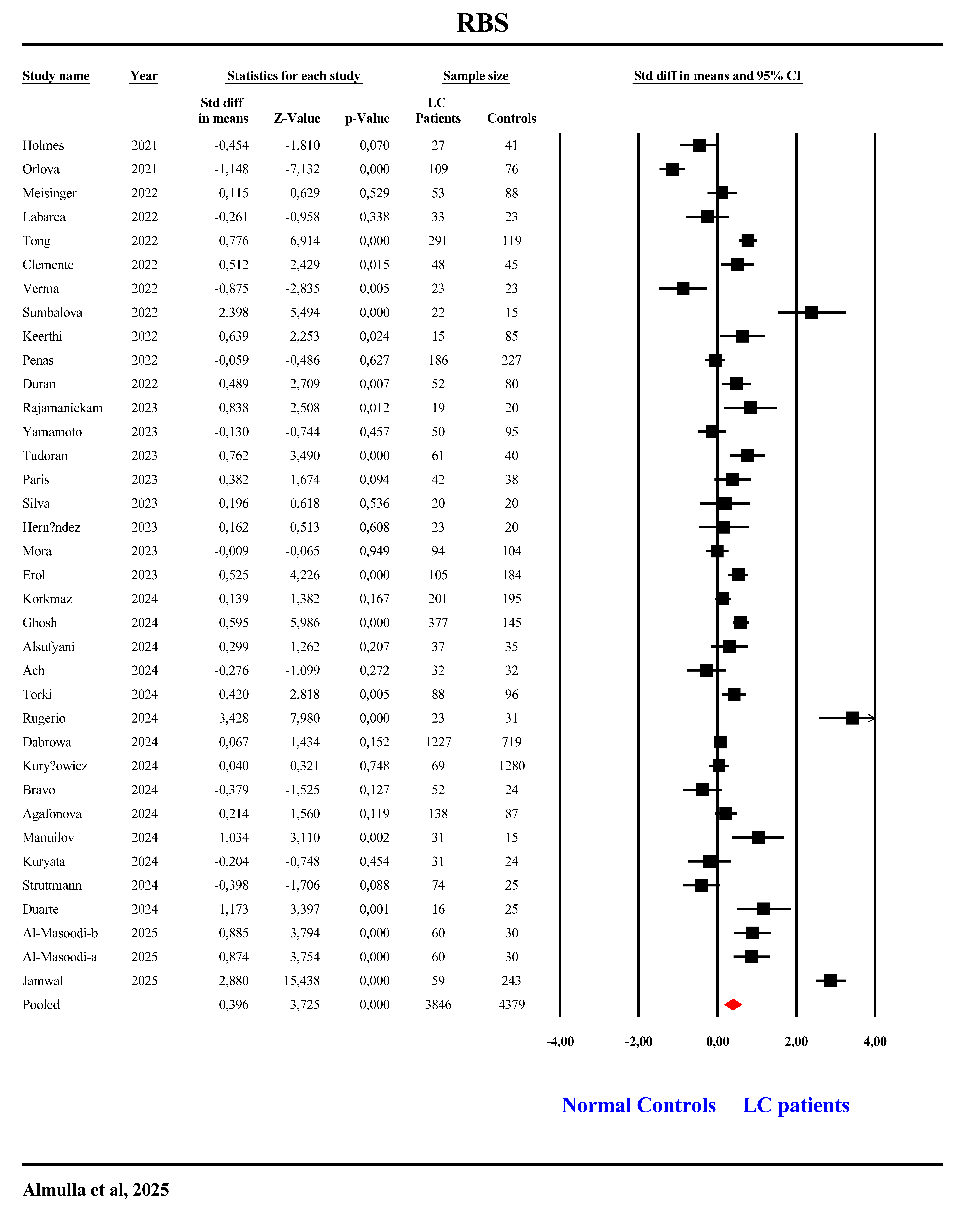

ESF, Figure 2. The forest plot of random blood sugar (RBS) in patients with Long COVID (LC) and normal controls.

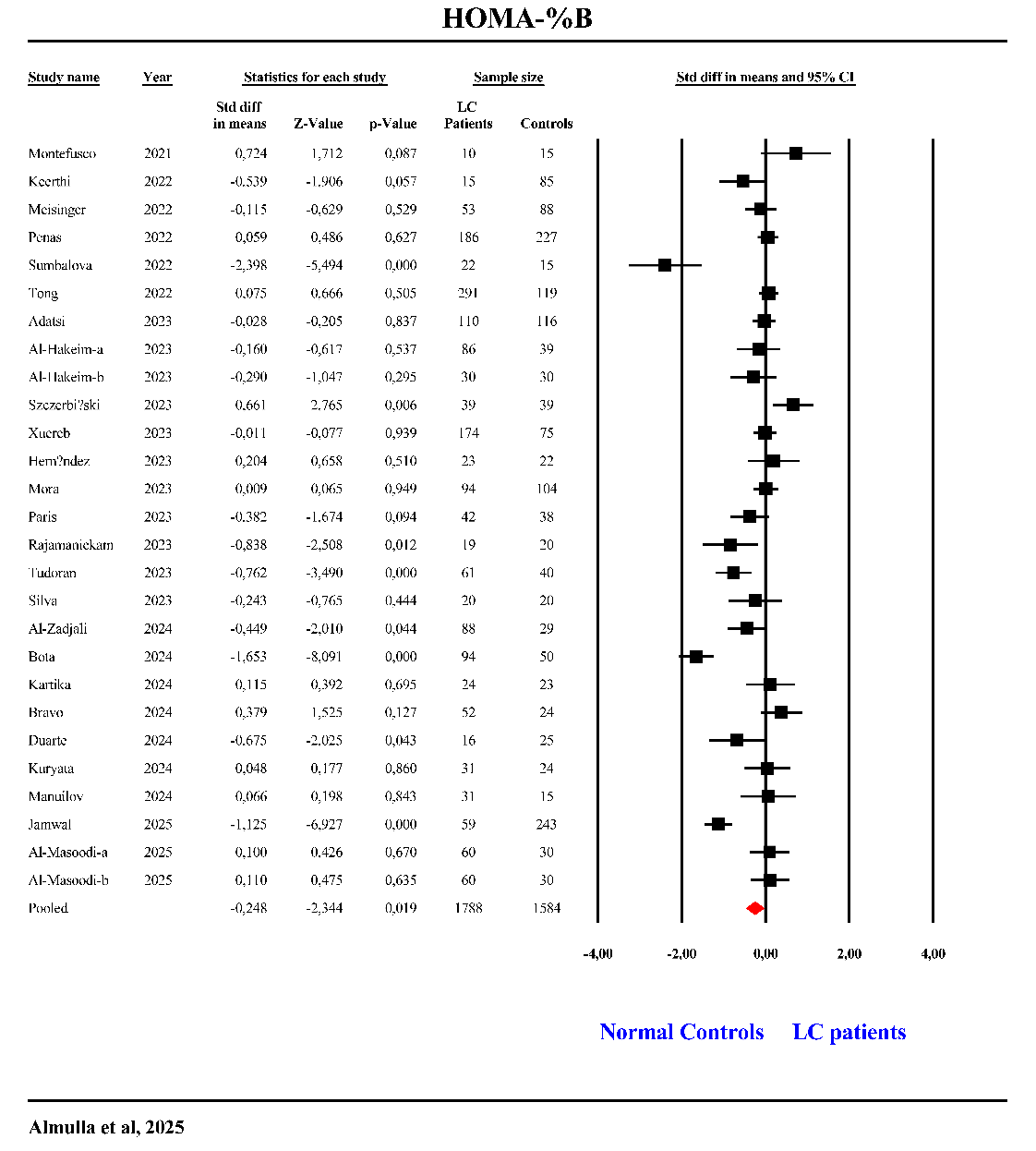

ESF, Figure 2. The forest plot of HOMA-%B in patients with Long COVID (LC) and normal controls.

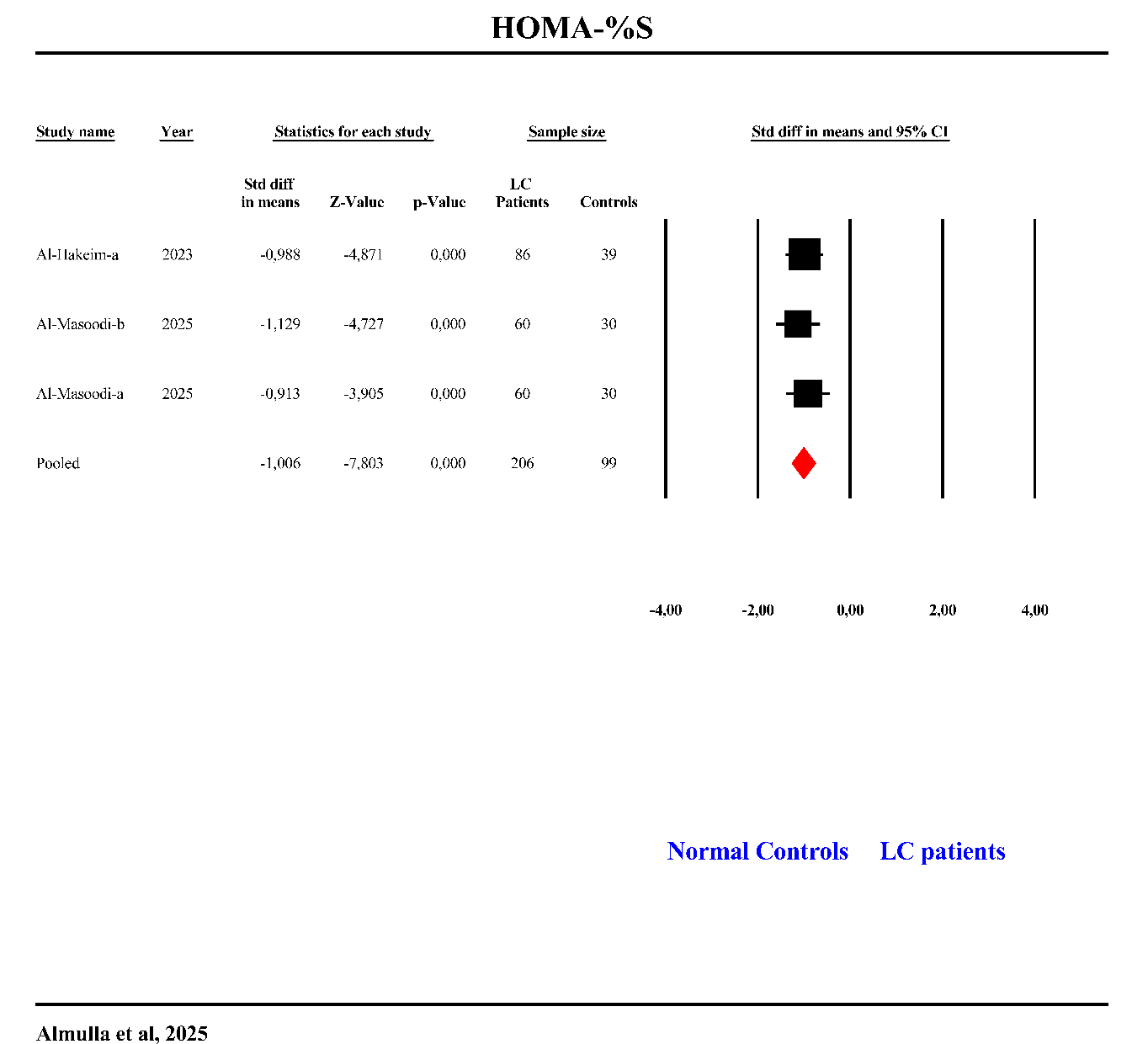

ESF, Figure 2. The forest plot of HOMA-%S in patients with Long COVID (LC) and normal controls.
